# Is education a risk factor for Parkinson’s disease?: A systematic review and meta-analysis

**DOI:** 10.1101/2025.07.02.25330781

**Authors:** Laurenz Lammer, Chris Patrick Pflanz, Megan Kirk, Ammar Shahabuddin, Sarah Bauermeister

## Abstract

Parkinson’s disease (PD) has shown a twofold increase in global prevalence over the past 25 years, making research into its causes pivotal. Curiously, higher educational attainment has often been linked to a higher risk of PD. Yet, to our knowledge, these findings have not been validated in a systematic literature review and meta-analysis.

Thus, we systematically searched PubMed, Psychinfo, Web of Science and Embase and performed citation and similar article searches. We included reports of non-interventional quantitative studies that assessed the association between education and the risk of PD. Furthermore, we included studies that evaluated the association between educational level and symptom burden. For the primary outcome, we performed a random-effects meta-analysis. The secondary outcome was synthesized qualitatively. We conducted multiple a priori subgroup analyses. Risk of bias was assessed using the QUIPS tool. This study was registered on PROSPERO (ID: CRD420250651033).

Of 23,648 identified reports, 36 papers met inclusion criteria. Most of these had moderate or high overall risk of bias. Meta-analytic findings yielded no significant association between education and risk of PD, with a pooled odds ratio of 1.09 (k = 24; 95% CI: 0.92 – 1.28). Sensitivity analyses yielded similar results. Subgroup analyses did not explain observed heterogeneity. There was no evidence of small study effects or p-hacking. Education appeared to be associated with reduced PD symptom severity but the number and quality of studies was limited. Education is unlikely to be an independent risk factor for PD. Study heterogeneity and methodological limitations preclude firm conclusions, though.

## Introduction

PD is the second most common neurodegenerative disease^1^ and affects around 12 million individuals globally^2^. Its incidence has surged over the past decades, far exceeding an increase explainable by demographic changes alone^3^. To date, no disease-modifying treatment for PD has been found^4,5^ and only around one third of cases can be attributed to mono- and polygenetic causes^6^.

Henceforth, it is a paramount aim of PD research to identify and understand the non-genetic causes of disease to facilitate the discovery of preventive interventions^6^. Yet, research on risk factors for the disease has not yielded any actionable results beyond recommendations of increased physical activity^7^. Instead, findings have repeatedly puzzled investigators. Systematic reviews and meta-analyses concerning smoking and alcohol consumption, behaviors almost invariably linked to worse health outcomes, identified them to be associated with lower risk of PD^7,8^.

Curiously, multiple studies also linked less education to a lower risk of PD^9–12^ which was corroborated by a recent mendelian randomization study^13^. Amongst neurological disorders, PD seems to be a unique case whose age-standardized prevalence increases with a higher regional socio-demographic index^3,14^. However, some studies failed to replicate the association of PD and education^15,16^ or had inconsistent ^17^ or contradictory results^18^.

To our best knowledge, no systematic review and meta-analysis has been conducted to investigate the association between education and PD risk. Hence, the evidence level is unsatisfactory. Therefore, the purpose of this systematic review was to critically assess the cumulative evidence regarding the association between formal educational attainment and the risk of PD. Specifically, we aimed to determine whether reported significant findings reflected an empirical relationship or were more likely to be spurious. To this end, we performed a comprehensive and systematic search for relevant studies and conducted a random-effects meta-analysis to quantitatively synthesize effect estimates. Additionally, we qualitatively synthesized the available evidence on the effect of educational attainment on PD symptom burden.

Furthermore, we also aimed to better understand the mechanism of action of the purported association. We hypothesized that the observed correlations actually stem from better access to medical care amongst highly educated people. Additionally, positive associations might be mediated by the negative correlations of education with smoking and the inverse link of smoking and the risk of PD. Therefore, we explored these hypotheses by adding multiple pre-specified subgroup analyses to our meta-analysis.

## Methods

### Protocols

This manuscript conforms to the Preferred Reporting Items for Systematic Reviews and Meta-Analyses (PRISMA) guidelines^19^. We registered this study with the International Prospective Register of Systematic Reviews (PROSPERO), number CRD420250651033. We amended the protocol of the registration to include a pre-specification of a meta-analysis. We had decided to perform a systematic quantitative analysis and discuss and explore the heterogeneity of our data rather than to leave the door open to unsystematic quantitative approaches such as vote counting^20^.

### Software and code

We used Rayyan^21^ and SRDR+^22^ for screening and data extraction, respectively. All calculations were performed in R (Version 4.4.3) and all code can be found on https://github.com/LaurenzLammer/pd_education_ma. The extracted data have been uploaded to the osf (https://osf.io/j9hp2). The analysis script on github is directly linked to the dataset in the osf repository so that all analyses and figures can be reproduced easily. Additionally, it produces many more plots and tables on our sensitivity analyses that did not make it into the appendix. Moreover, the scripts can be downloaded and modified to allow the exploration of the effects of changing assumptions made in our calculations (e.g. the correlation when aggregating effect sizes).

### Study selection

#### Population

We included studies of human adults (>= 18a) that had at least 20% of participants above the age of 50 years due to PD’s age dependence. We considered original research studies of individual participants such as cohort studies, cross-sectional studies, case-control studies and studies using instrumental variable methods (e.g. Mendelian Randomization) as well as studies investigating predictors of symptom severity in clinical samples but excluded all other forms of studies (e.g. qualitative studies, case reports, ecological studies or intervention studies).

#### Exposure

We only included studies that investigated education as a predictor of interest. Studies that merely assessed education as descriptive statistic or a control variable were not included.

#### Outcome

Our primary outcome was the risk of idiopathic PD. Studies were included independent of whether they measured this as an odds ratio (OR), risk ratio (RR), hazard ratio (HR) or otherwise. Studies on atypical parkinsonian syndromes (e.g. multiple system atrophy) did not meet our inclusion criteria. Our secondary outcome was the severity of PD’s core symptoms (tremor, rigor, akinesia/bradykinesia, postural instability). Reports had to be either in English or German to be included in the review to enable dual screening and dual data extraction.

### Search strategies

Fig. S1 provides an overview of our search and selection strategy. We searched PubMed and Web of Science (WoS) directly, and PsychInfo and Embase independently via Ovid for relevant records using a combination of relevant keywords and controlled vocabulary without restrictions on publication date. Our search strategy was developed based on previous work on aetiological search filters^23^. We piloted the Pubmed search strategy using a set of nine studies identified in an unsystematic literature search and performed a peer review of electronic search strategies (PRESS) for all strategies^24^. The appendix contains full search terms and dates of the database searches. Additionally, we iteratively performed backward citation searching (WoS), forward citation searching (both WoS and Scopus) and similar articles searching (PubMed) on all records included after full-text screening in April 2025. MK & LL manually deduplicated potential duplicates identified by Rayyan. The PRISMA flowchart^19^ in Fig 1 provides information on the number of records identified by the different components of the search strategy.

**Figure 1:**
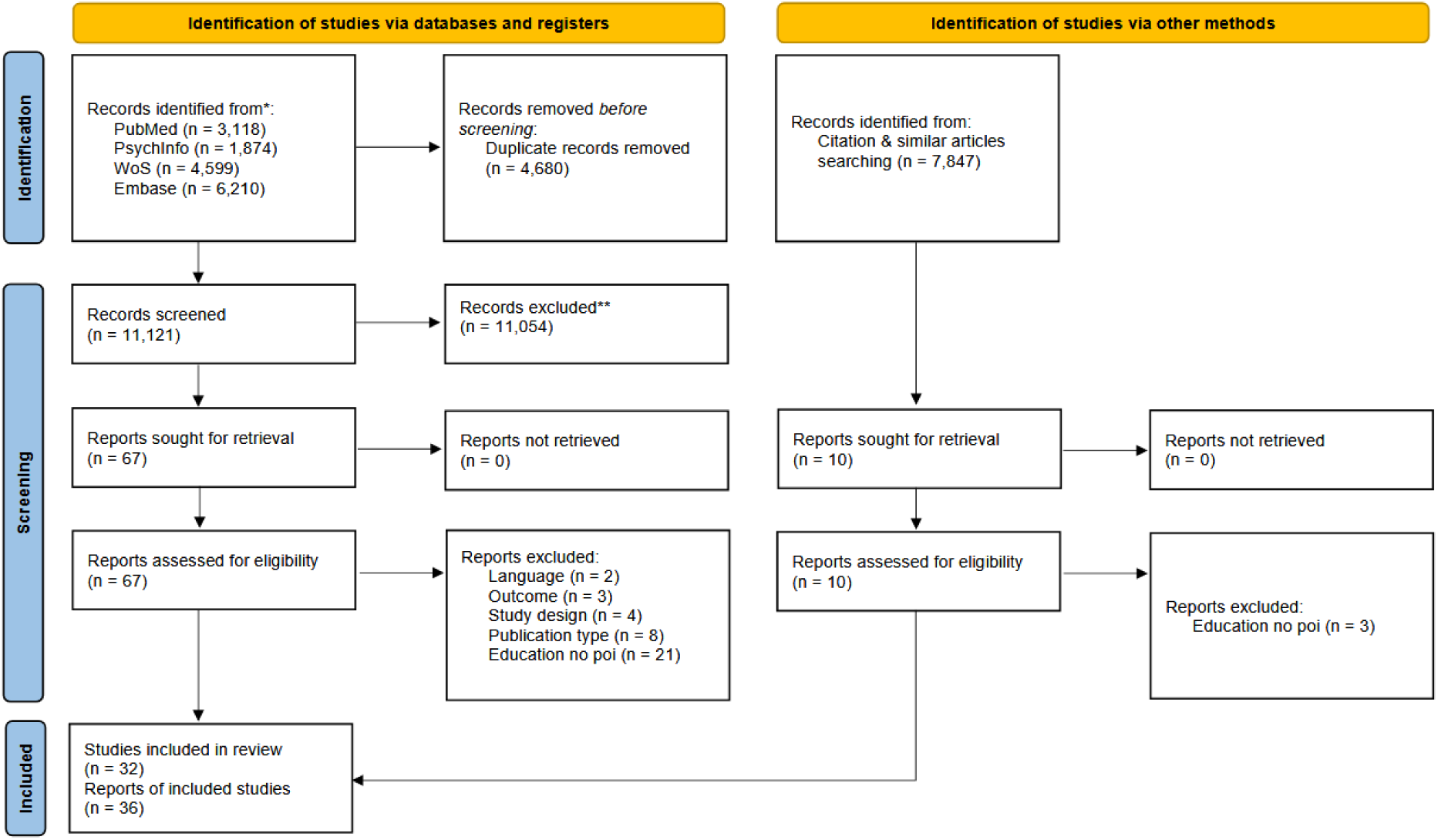
Flowchart delineating the search and screening process according to the 2020 PRISMA statement. Education no poi = Education was not investigated as a predictor of interest.

### Screening

MK and LL independently performed title and abstract screening on 50% of the records identified in the database searches after deduplication while being masked to each other’s decisions. LL screened the remaining records. In case of uncertainty or disagreements, the screeners discussed the records in question until they reached a consensus.

MK, PP and LL did the independent and masked full-text screening of all remaining records. Each record was screened by at least two reviewers (LL screened all). Consensus on inclusion / exclusion in case of doubt or disagreement was reached by a discussion in the team.

### Retractions and errata

We searched for retractions or corrections of reports selected after full text screening using the retraction watch database on April 24^th^ 2025.

### Data extraction

We designed a data extraction form in SRDR+ based on the Cochrane Handbook ^25^, which two review authors (AS and LL) used to independently extract data from eligible studies. We compared the extractions using SRDR+’s comparison tool and resolved any discrepancies through discussion. Please refer to the appendix for a list of extracted information.

After consolidating the extractions, we manually adapted the data exported from SRDR+ to align them with the format required for meta-analysis in R.

### Risk of bias assessment

For every included study the risk of bias (RoB) was determined using the revised QUIPS tool^26^ by the two extractors independently. Assessors were not masked to any aspects of the reports when making RoB assessments^27^ but to each other’s judgements. Any conflicts were resolved through discussion between the two. For the assessment of the overall RoB of a study, the worst score across the most important QUIPS domains (education measurement, PD measurement, confounding, analysis and reporting) was decisive. We created a traffic light RoB plot for the primary and secondary outcome using the rob_traffic_light function from **{robvis}**. Additionally, we created weighted domain-summary RoB plots using the rob_summary function of **{robvis}** to illustrate overall RoB per domain. For the secondary outcome, we created an unweighted plot.

### Linking multiple reports of the same studies

If multiple reports based on the same studies were identified, we prespecified to choose the report with the lowest RoB according to the QUIPS tool for quantitative synthesis.

### Data synthesis

We categorized the identified studies depending on whether they investigated our primary (risk of PD) or secondary outcome (severity of core PD symptoms). Data were considered suitable for pooling if participants’ risk of PD was investigated in relation to their level of formal education.

Prior to analysis, we log-transformed ORs. To obtain a log-transformed standard error for each effect, we applied the SE.SMD_from_OR.CI function from **{metaHelper}** to the upper and lower limits of the OR confidence interval and multiplied the result by 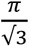 (*Chinn*, 2000).

As education across time and space as well as the study designs are highly heterogenous, we calculated a random-effects model to pool effect sizes. We employed the metagen function of **{meta}** and used the Paule-Mandel procedure ^29^ to calculate the heterogeneity variance τ^2^ and the Knapp-Hartung adjustment ^30^ to calculate the confidence interval around the pooled effect.

#### Assessment of heterogeneity and subgroup analyses

We tested the statistical heterogeneity with the Chi^2^ test of Cochran’s Q at a significance level of 0.05 and the I^2^ statistic as returned by the metagen function. I^2^ values of 25%, 50% and 75% were considered indications of low, moderate and substantial heterogeneity, respectively^31^.

We prespecified that we would investigate outliers and influential cases if I^2^ exceeded 50%. As a sensitivity analysis, we ran the find.outliers function from **{dmetar}** to identify studies whose effect sizes do not overlap with the pooled effect’s confidence interval and recalculated our analysis excluding these studies. To investigate influential cases, we employed the InfluenceAnalysis function from **{dmetar}**.

In our registration, we prespecified three study characteristics for subgroup analyses to explain between-study heterogeneity. As education is negatively correlated with smoking and smoking is negatively correlated with the risk of Parkinson’s disease^7^, we conducted a subgroup analysis to test whether controlling for smoking or not contributed to the heterogeneity. To this end, we updated the metagen meta-analysis setting the argument “subgroup” to the variable distinguishing studies by this characteristic.

Furthermore, we distinguished studies that assessed the general population and those that did not (e.g. inpatient samples) and tested if this explains some of the heterogeneity. Likewise, we differentiated studies that clinically examined all participants and those that did not and investigated this classification’s effect on heterogeneity. Moreover, we explored if effects differ between high- and, low- and middle-income countries by calculating another subgroup analysis. Please see the appendix for details on the categorization.

#### Sensitivity analysis

We conducted sensitivity analyses calculating meta-analsyses only including a) studies that reported ORs (no RRs, HRs, etc.), b) no outliers as identified by find.outliers, c) no studies deemed to have an overall high RoB, d) no mendelian randomization studies, e) only the Swedish register-based study with the lowest RoB.

### Assessing small study effects, publication bias and p-hacking

As recommended by Harrer et al.^32^ we applied a set of methods to investigate publication bias, small study effects and p-hacking. We provide a standard funnel plot produced by the funnel function from **{meta}** to allow for a visual examination of small study effects. We had prespecified that we would perform all following methods on a subset of studies after the exclusion of outliers detected by find.outliers if I^2^ exceeds 75%.

We performed Rücker’s limit meta-analysis method using the limitmeta function from **{metasens}** and produced the adapted funnel plot using the funnel.limitmeta function. Additionally, we performed a p-curve analysis using the pcurve function of **{dmetar}**. Furthermore, we used the pcurve function to calculate a corrected effect size by providing a sample size for every study.

### Effect measures

Most of the studies used ORs as effect measures. If adjusted and unadjusted ORs were reported, we used the adjusted ones. We treated RRs as ORs if the sample was population based and hence the base rate of PD was low. There is no clear guidance on how to handle HRs or more infrequent outcome measures like standardized incidence ratios ^20,33^. For the sake of completeness, we treated them as ORs, too but ran a sensitivity analysis only including studies reporting ORs. We handled ORs from mendelian randomization studies analogously.

We aligned the direction of effects so that higher ORs indicate higher odds amongst more educated individuals. We adapted reported effect sizes from reports that did not base their estimates on a group comparison but e.g. years of schooling to homogenize them with the others. If reports included effect estimates for multiple comparisons (e.g. illiterates vs others and <5 years of schooling vs others) or Mendelian Randomization studies provided effect size estimates based on various calculation methods, we aggregated the estimates using the aggregate function of **{metafor}** setting ρ (the correlation of the sampling errors) to 0.6. In two cases we calculated an OR using oddsratio() from **{epitools}** based on raw count data. If an OR was reported without a standard error (SE), or no OR but beta-coefficients and SEs from logistic regression were reported, we used standard formulae to calculate the effect size with its confidence interval. Please see the appendix for details on all these procedures.

### Certainty of evidence assessment

We applied the GRADE approach to certainty of evidence assessment^34,35^. All factors determining the certainty of evidence were evaluated according to the GRADE handbook^36^. However, as suggested by Morgan et al. and Foroutan et al., we did not downgrade observational evidence on principle as the best evidence for our research question is from observational studies^37,38^. We did not translate QUIPS categories to GRADE categories but followed the advice of the GRADE handbook and attempted to judiciously consider the contribution of each study to the overall results^36^. The included studies did not allow us to discern a baseline risk of Parkinson’s disease from the body of evidence. Hence, we relied on the analysis of Elbaz et al. that calculated a lifetime risk of 1.65%^39^ as an approximation. It should be noted that other analyses produced substantially higher estimates, though^40^. We calculated the absolute risk as p_high education_ = (OR * p_baseline_) / (1 – p_baseline_ + OR * p_baseline_)^41^ and derived upper and lower limits analogously.

## Results

### Study selection

We identified 15,801 records across the four databases. After deduplication, we screened 11,121 records, from which we reviewed 67 full-text documents, and finally included 29 papers. Citation and similar articles searches yielded an additional 7,847 records of which 10 were chosen for full-text screening and 7 were ultimately included in our analysis, resulting in a total number of 36 reports included after full-text screening. At title and abstract screening, the agreement was 99.39% and Cohen’s kappa 51.77% because of the high chance of random agreement to exclude. At full-text screening, the agreement between the three raters was 90.3 % and 90.2% and Cohen’s kappas were 80.5% and 80.5%. Fig. S1 provides details on the screening process. A table on characteristics of excluded studies^42–50^ with initial disagreement or uncertainty at full-text screening is provided in the appendix (Table S1)^51^. None of our records were identified in the Retraction Watch database.

### Description of studies

#### Characteristics and results of individual studies

The identified studies employed a broad range of approaches to investigate the link between education and the risk of PD. Multiple studies on our primary outcome used data from pre-existing epidemiological studies like the UK Biobank (n = 10, 36%), five studies were based on large health registers from Scandinavia or China (18%) and others purposively sampled participants for classic case-control designs (n = 9, 32%). Mendelian randomization studies based their estimates on large GWAS (n = 4, 14%). All studies on our secondary outcome used patient samples. The different study designs went hand-in-hand with divergent approaches to the assessment of PD. Only a minority of reports on our primary outcome assessed all participants based on established diagnostic criteria (n = 9, 32%). The operationalization of education was highly heterogenous, too. Across both outcomes, seven studies assessed it continuously (19%), while others used a dichotomous (n = 9, 36%) or polytomous categorization (n = 20, 56%) with diverse cut-offs. Moreover, while ORs (n = 21, 75%) were the most prevalent type of effect estimate for our primary outcome, five reports calculated HRs (18%) or yet other effect measures (n = 2, 7%). Furthermore, while most studies controlled for the effects of age and gender (n = 26, 72%), the additionally included confounders were highly heterogenous between studies. Several studies were conducted in low- or middle-income countries (n = 10, 28%) but the majority of reports were from high-income countries (n = 26, 72%). We excluded one of the 36 reports from further analysis due to redundance^52^. We found 27 reports investigating our primary outcome^11,12,15–17,53–74^ and 7 for our secondary outcome^49,75–81^. One study using the age at symptom onset^80^ as its outcome did not fit in either group exactly and we grouped it with the studies on our secondary outcome. Table 1 provides an overview of the reports on our primary outcome included in our review. The similar Table S2 contains information on reports on our secondary outcome.

**Table 1.** Characteristics of the Included studies for the primary outcome.

| Characteristics of the included studies for the primary outcome |  |  |  |  |  |  |  |  |  |  |
| --- | --- | --- | --- | --- | --- | --- | --- | --- | --- | --- |
| Report | Study design | Location | Sample size | Diagnostic criteria | Representative sample | All participants examined | Controlled for smoking | Income category | Effect measure | Effect |
| Achbani 2022 | case-control | Morocco | 540 |  | no | no | no | LMIC | OR | none vs university: 0.5 (0.21 - 1.02)<br>primary vs university: 0.51 (0.23 - 1.1)<br>secondary vs university: 0.56 (0.25 - 1.28) |
| Baldereschi 2003 | cross-sectional | Italy | 4,496 | study specific | yes | yes | yes | HIC | OR | per year: 0.86 (0.66 - 1.1) |
| Chen 2015 | longitudinal | China | 74,941 |  | yes | no | no | LMIC | OR | =< primary vs < high school: 1.3 (0.7 - 2.4)<br>=< primary vs >= high school: 1.6 (1 - 2.7) |
| Fardell 2020 | case-control | Sweden | 1,189,134 | ICD | yes | no | no | HIC | HR | no highschool vs highschool: 1.2 (1.01 - 1.32)<br>no highschool vs university: 1.37 (1.19 - 1.58) |
| Frigerio 2005 | case-control | USA | 392 | study specific | yes | yes | no | HIC | OR | =<9 vs >9a: 1.8 (1 - 3.3) |
| Gialluisi 2023 | longitudinal | Italy | 23,901 | ICD | yes | no | no | HIC | HR | primary vs lower secondary: 0.76 (0.5 - 1.15)<br>primary vs post secondary: 0.69 (0.38 - 1.25)<br>primary vs upper secondary: 1.03 (0.72 - 1.49) |
| Gupta 2014 | case-control | India | 194 | UK PDS BB | no | yes | no | LMIC | OR | illiterate vs others: 0.92 (0.52 - 1.62) |
| Huang 2023 | instrumental variable | European ancestry | 482,730 |  | yes | no | yes | HIC | OR | IVW: 1.38 (1.08 - 1.75)<br>EGGER: 1.76 (0.67 - 4.65)<br>WM: 1.26 (0.92 - 1.75)<br>S Mode: 0.74 (0.28 - 1.93)<br>W Mode: 1.04 (0.44 - 2.42) |
| Jacobs 2020 | longitudinal | UK | 501,682 |  | yes | no | no | HIC | OR | per year: 0.95 (0.82 - 1.1) |
| Kuopio 1999 | case-control | Finland | 369 |  | no | yes | no | HIC | OR | primary vs < upper secondary: 0.89 (0.55 - 1.42)<br>primary vs >= upper secondary: 1.04 (0.5 - 2.19)<br><10a vs >= 13a: 0.98 (0.42 - 2.27)<br>11-12a vs >= 13a: 1.16 (0.59 - 2.27) |
| Kyrozis 2013 | longitudinal | Greece | 25,407 |  | yes | no | yes | HIC | HR | per 3 years: 0.81 (0.57 - 1.15) |
| Li 2009 | longitudinal | Sweden |  |  | no | no | no | HIC | SIR | <9 vs other men: 0.96 (0.94 - 0.99)<br><9 vs other women: 1.04 (1 - 1.08) |
| Li 2024 | instrumental variable | European ancestry | 482,730 |  | yes | no | yes | HIC | OR | IVW: 1.47 (1.21 - 1.78)<br>WM: 1.46 (1.07 - 1.98)<br>EGGER: 1.66 (0.85 - 3.22) |
| Li 2025 | longitudinal | UK | 452,492 |  | yes | no | no | HIC | HR | =< A-Levels vs professional/academic degree: 0.98 (0.92 - 1.04) |
| Llibre-Guerra 2022 | cross-sectional | Latin America | 12,734 | UK PDS BB | yes | yes | no | LMIC | PR | per level (of 5)*: 1.39 (1.03 - 1.89) |
| Martyn 1995 | case-control | UK | 515 |  | no | no | no | HIC | OR | Left school <14a vs left school >14a: 0.8 (0.52 - 1.35)<br>Left school <14a vs further education: 0.8 (0.47 - 1.55) |
| Najafi 2023 | case-control | Iran | 1,523 | MDS | no | yes | no | LMIC | OR | =< primary vs secondary: 1.02 (0.76 - 1.35)<br>=< primary vs academic: 2.17 (1.58 - 2.99) |
| Osler 2022 | longitudinal | Denmark | 656,751 | ICD | yes | no | no | HIC | HR | 7-9a vs 10-11a: 1.03 (0.97 - 1.1)<br>7-9a vs >=12a: 1.24 (1.14 - 1.35) |
| Rocca 1996 | case-control | Italy | 186 | study specific | yes | yes | no | HIC | OR | illiterates vs others: 1.9 (0.9 - 4.4)<br><5a vs others: 1.1 (0.6 - 2.2) |
| Shi 2022 | instrumental variable | European ancestry | 482,730 |  | yes | no | yes | HIC | OR | IVW: 1.24 (1.02 - 1.51)<br>EGGER: 1.45 (0.68 - 3.09) |
| Tan 2007 | case-control | China | 314 | other | no | yes | no | LMIC | OR | =< junior middle vs >= senior middle: 1.63 (0.98 - 2.71) |
| Taylor 1999 | case-control | USA | 287 | Ward & Gibb | no | no | no | HIC | OR | per year: 0.31 (0.18 - 0.53) |
| Torti 2020 | case-control | Italy | 1,166 | UK PDS BB | no | no | yes | HIC | OR | primary vs middle: 0.75 (0.51 - 1.1)<br>primary vs high school: 0.87 (0.6 - 1.26)<br>primary vs academic: 0.98 (0.65 - 1.48) |
| Wijeyekoon 2017 | case-control | Sri Lanka | 246 | UK PDS BB | no | no | yes | LMIC | OR | < A-Levels vs >= A-Levels: 1.92 (0.86 - 4.27) |
| Wirdefeldt 2005 | case-control | Sweden | 2,856 |  | yes | no | yes | HIC | OR | compulsory vs > compulsory - external controls: 1.35 (1.05 - 1.75)<br>compulsory vs > compulsory - co-twin controls: 0.94 (0.58 - 1.53) |
| Zhang 2022 | instrumental variable | European ancestry | 525,556 |  | yes | no | yes | HIC | OR | IVW: 1.68 (1.07 - 2.62)<br>WM: 1.56 (0.83 - 2.94)<br>EGGER: 0.24 (0.04 - 1.54) |
| Zou 2015 | case-control | China | 9,493 | UK PDS BB | yes | no | no | LMIC | OR | <7a vs >= 7a: 1.07 (0.82 - 1.41) |

ICD = International Classification of Diseases; UK PDS BB = UK Parkinson's Disease Society Brain Bank; MDS = Movement Disorder Society; Ward & Gibb = Research diagnostic criteria for Parkinson's disease by Ward & Gibb 1990; OR = odds ratio; HR = hazard ratio; SIR = standardized incidence ratio; PR = prevalence ratio; IVW = inverse variance weighted; WM = weighted median; S Mode = simple mode; W Mode = weighted mode; \* = no education, did not complete primary, completed primary, secondary, or tertiary education

#### Risk of bias

Most studies (86%) were deemed to be at moderate (n = 15, 43%) or high (n=15, 43%) overall RoB, with only five studies (14%) showing low overall RoB. Each study’s domain-specific and overall RoB is tabulated in Table S3 for our main outcome and in Table S4 for our secondary outcome. Figs S2 and S3 show a traffic light plot and a domain-summary RoB plot for our primary outcome, respectively. Please refer to Fig S4 for plots including removed linked studies and to Figs S5-6 for plots for our secondary outcome.

### Results of syntheses

#### Quantitative synthesis of education and the risk of Parkinson’s disease

Prior to quantitative synthesis, we linked reports on the same studies and chose the paper with the lowest RoB. Please see the appendix for details on linked reports.

We incorporated 24 studies that investigated the link between educational attainment and the risk of PD in our meta-analysis. Higher education was not significantly associated with higher odds of PD (OR (95% CI) = 1.09 (0.92 – 1.28); p = 0.294; I^2^ = 79.5%; absolute risk = 0.14% higher lifetime risk (-0.12% to +0.45%). The heterogeneity between studies was high and included both studies with odds ratios significantly lower and significantly higher than 1. Details are provided in Table 2 and Fig 2 depicts a standard forest plot. Additionally, Fig S7 shows a drapery plot as an alternative illustration of individual studies and the main effect.

**Table 2.**
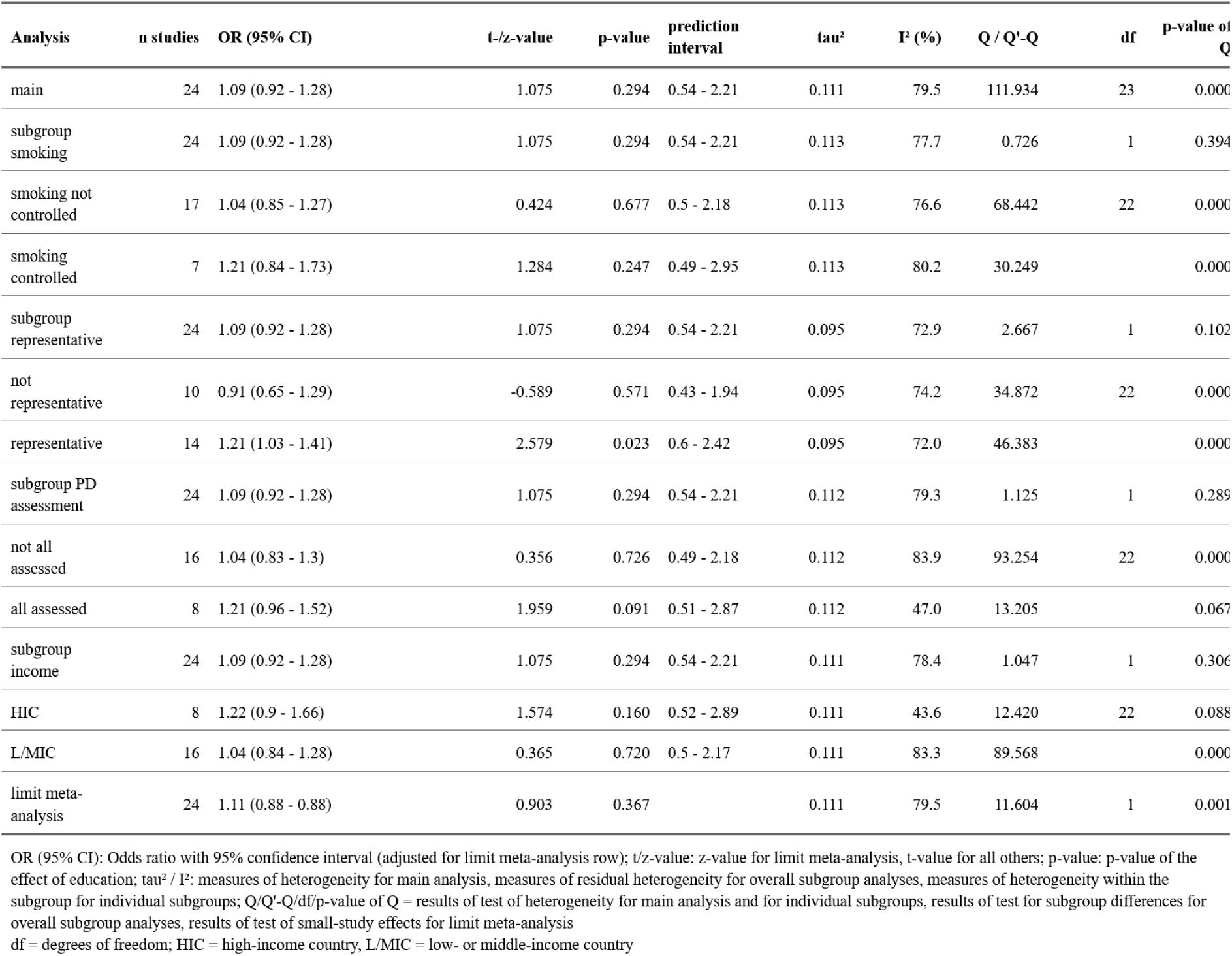
Results of meta-analyses of all studies.

**Figure 2:**
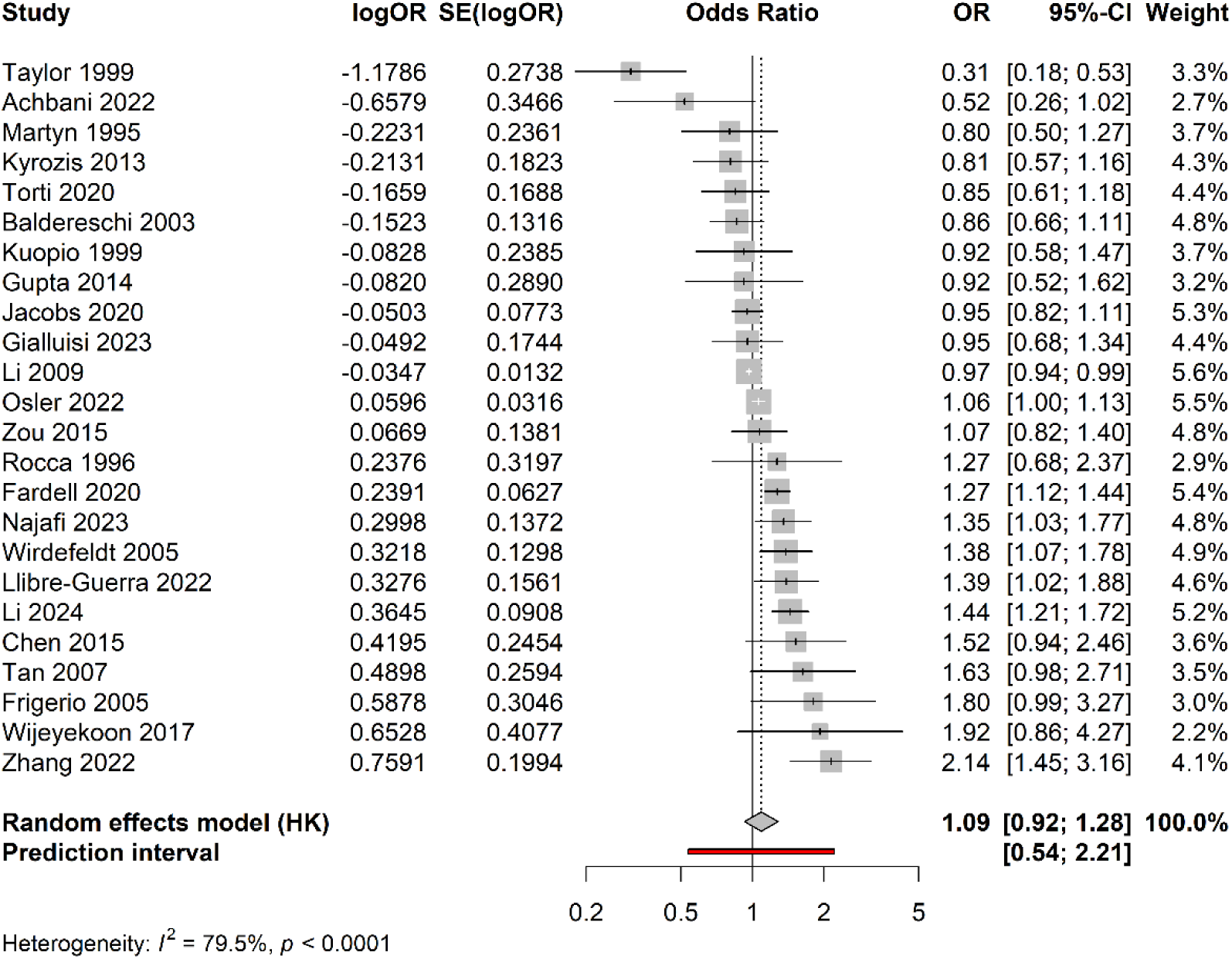
Forest plot of our main meta-analysis.

### Subgroup analyses

None of the following factors explained a significant share of the heterogeneity: whether studies controlled for smoking, used a representative sample, or assessed all participants. Neither did comparing studies from high- to low- or middle-income countries. Results for each of the subgroup analyses and the respective subgroups are presented in Table 2.

### Outliers and influential cases

We detected two outliers (one with a low and one with a high OR). Excluding these two studies did not affect the effect size in a meaningful way but the heterogeneity was reduced slightly (OR (95% CI) = 1.1 (0.98 - 1.24); p = 0.089; I^2^ = 73.4%). The subgroup analyses did not differ substantially in this subsample of studies either. Further information can be found in Table S5 and a forest plot is provided in Fig S8.

This is corroborated by a Baujat plot (Fig S9) that also identified these two studies as contributing heavily to the heterogeneity but not to the overall effect size. No study in the Baujat plot contributed heavily to both the heterogeneity and overall effect size. None of our further influence tests (externally standardized residuals, DFFITS value, Cook’s distance, Covariance ratio, leave-one-out τ^2^ and Q-values, hat value, and study weight) identified any study as particularly influential (Fig S10). Furthermore, 95% confidence intervals of leave-one-out meta-analyses consistently included 1 (Figs S11-12).

### Sensitivity analyses

None of our additional sensitivity analyses (excluding mendelian randomization studies, excluding studies with a high overall RoB, excluding studies reporting effect measures other than OR, including only one Swedish register-based study) led to meaningful differences in either heterogeneity or effect size in the main analysis or the subgroup analyses (Tables S6-9 and Figs 13-16).

#### Qualitative synthesis of education and Parkinson’s disease severity

We identified seven studies that investigated differences in PD patients symptom severity related to education. Only one study that assessed the Unified Parkinson’s Disease Rating Scale (UPDRS) motor subscale did not find a significant effect of education^76^ while five others (two of them with overlapping samples, one including non-idiopathic forms of parkinsonism) found significant associations of higher education with better motor performance^75,77–79,82^. In addition, the former study did find a significantly better performance on the Berg Balance scale amongst more highly educated patients. Furthermore, gait speed was positively linked to education during verbal dual-tasking but not in four other conditions in a Spanish study^81^. Moreover, a longitudinal study of PD patients identified a significantly higher hazard ratio of developing levodopa-induced dyskinesia amongst patients with lower education^78^.

### Reporting biases

Fig 3 shows a standard funnel plot of the studies included in our meta-analysis that highlights outliers but does not depict major asymmetry. Due to the high heterogeneity in our main meta-analysis, we performed Rücker’s limit meta-analysis and a p-curve analysis on a subset of studies after excluding outliers. The former’s test for small study effects was significant (p = 0.001) but it also indicated substantial residual heterogeneity beyond small-study effects and the unadjusted and adjusted effect estimates were practically indistinguishable apart from the wider confidence interval of the latter (please see Table 2 for details). A funnel plot showing the shrunken effect sizes is depicted in Fig S17. The p-curve analysis did not find evidence of p-hacking at an estimated power of 59% (test for flatness: p_full_ = 0.829, p_half_ = 0.999) and indicated the presence of evidence for an actual effect (test for right-skewness: p_full_ = 0.003, p_half_ = 0.002) but this must be disregarded as the significant p-values stem from studies both indicating significantly higher and significantly lower risk (please also see Fig. S18 and Table S10). The p-curve-based estimate of effect size d = 0.008 is miniscule and must be deemed unreliable due to the heterogeneity being >50% even after excluding outliers.

**Figure 3:**
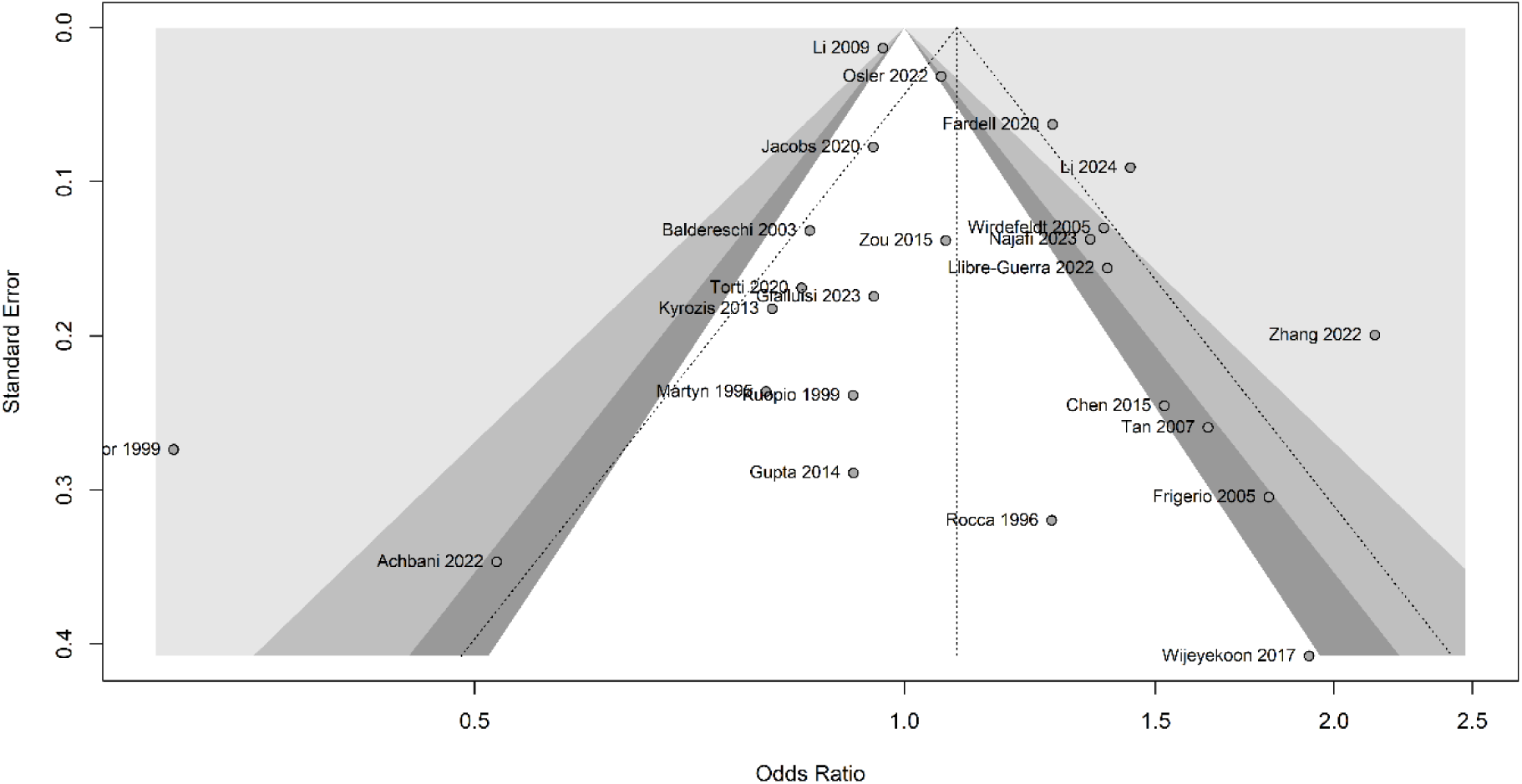
Funnel plot of the studies included in our meta-analysis. The dotted lines are centered around the overall effect size. The successively lighter grey funnels correspond to significance levels of 0.1, 0.05 and 0.01 for the individual studies.

### Certainty of evidence

Table S11 depicts the GRADE evidence profile for our primary and secondary outcome.

#### Education and the risk of Parkinson’s disease

As we applied a random-effects meta-analysis, the contribution of each study to the overall certainty of evidence is similar. The best evidence for the effect of educational attainment on the risk of PD is from observational studies. Thus, the design of the included studies does not merit downgrading the certainty of evidence^83,84^ and the a priori certainty of evidence is high. Concerns regarding confounding, clarity of eligibility criteria and outcome measurement in some of the studies should result in a downgrade by one level for risk of bias, though. Furthermore, the large inconsistency in results, that we could not explain by subgroup analyses, led us to downgrade the certainty by two additional levels. The directness of evidence, the availability of very large and precise studies, and the funnel plot symmetry as well as the results of the limit meta-analysis and p-curve spared us from further downgrading for imprecision or publication bias.

However, the absence of a large magnitude of effect and a dose-response gradient preclude an increase of the certainty of evidence. Plausible residual confounding is not a reason for greater certainty of evidence, either. Hence, the risk of bias and the inconsistency of results caused us to judge the overall certainty of evidence to be **very low**.

#### Education and the risk of Parkinson’s disease

For our secondary outcome, most judgements are analogous. As the results across studies were rather homogenous, we did not have to downgrade for inconsistency, though. However, the smaller sample sizes and lower number of studies led us to downgrade by one level for imprecision. Thus, the risk of bias and imprecision resulted in an overall judgement of a **low** certainty of evidence for our secondary outcome.

## Discussion

This was the first known systematic review and meta-analysis to quantify the association between education and risk of PD. Overall, 36 papers were synthesized and included in this review. The results of our meta-analysis indicate that educational attainment is not linked to the risk of PD in either way. However, the moderate to high RoB of most studies and the high heterogeneity in effect sizes bar us from ruling out that there might be certain circumstances under which there is a positive or negative association. None of the theoretically plausible subgroup analyses revealed such a circumstance, though.

Concerning our secondary outcome, the scarcity of studies and their inconsistent risk of bias preclude definitive conclusions. However, the overall evidence points towards a protective effect of higher education against more severe PD symptoms.

### Further evidence

Several reports (e.g. conference abstracts) did not meet our inclusion criteria but should nonetheless be discussed here briefly. As in our meta-analysis, their results are highly heterogenous. Three reports found significantly higher education amongst cases when compared to controls^46,48,85^. Additionally, Yang et al. 2014 found a lower PD incidence in persons with low socioeconomic status^50^ and two mendelian randomization studies reported a positive association of IQ and PD risk^86,87^. However, four other studies, did not find significant differences by educational attainment^42,43,45,88^ and two reported significantly higher education in controls compared to cases^47,89^. Moreover, a study using dopamine transporter SPECT did not find significant differences by education level in tolerance against dopamine loss in PD^90^. In contrast, reports that did not meet our inclusion criteria that are relevant to our secondary outcome are well aligned. Lirani-Silva et al. 2017, Sohn et al. 2016 and Kotagal et al. 2015 all found worse motor symptoms amongst PD patients with lower educational attainment^49,79,91^. Some of these reports are linked to included studies, though.

### Limitations

We screened 23,648 reports and thus conducted a very extensive search of the literature. Yet, we cannot rule out that some relevant evidence might still have been missed due to location or language bias. However, these biases tend to reduce effect sizes and as we did not find an effect in either direction, this is less of a concern than in other circumstances. Another potential limitation of the review is that we could not perform dual title and abstract screening for all reports due to resource constraints. We did complete dual screening and extractions for all other stages, though and for over 5000 citations at title and abstract screening with an agreement of over 99% which makes us confident that the reliability of our findings is not hampered by this.

### Conclusions and outlook

Education is thought to influence health through various pathways. A key factor seems to be higher income with the resulting improved living conditions and better access to healthcare. Furthermore, safer working environments, salubrious behaviours and decisions, and beneficial effects of societal status and social networks are all likely to contribute to the positive effect of education on health^92,93^. However, all these mechanisms are believed to contribute to better health and hence cannot explain a higher risk of PD amongst more educated individuals.

Most studies that provided a hypothesis on why education was positively linked to risk of PD in their analyses attributed the association to increased health literacy and/or better access to healthcare amongst more highly educated individuals leading to more undiagnosed cases of PD amongst persons with lower educational attainment^15,72,80^. This is similar to the hypothesis that the lower prevalence of PD diagnoses in Blacks compared to Whites in the USA, despite no difference in Lewy body burden^94^, could be explained by disparities in access to health care^1,95^. The results for our secondary outcome are compatible with this notion, too. The greater symptom severity amongst patients with lower education could result from receiving a diagnosis only after more pronounced disease progression. Furthermore, this hypothesis is indirectly supported by evidence showing higher risk of PD in healthcare professionals^12,39,96^, who would be expected to have exceptional health literacy, and increased diagnostic latency amongst patients without health insurance^97^. Furthermore, the general underdiagnosis of PD elucidated by the high rates of new diagnoses in door-to-door prevalence studies^98,99^ adds to its plausibility.

However, there is no high-quality evidence on an association of education and diagnostic latency and all available studies did not find a significant link^100–103^. To test for quantitative evidence for this hypothesis, we aimed to explain some of the heterogeneity in effect sizes by categorizing studies in line with it. These subgroup analyses did not yield evidence supporting this hypothesis, though. However, we could only investigate this in a rather indirect manner and this must thus not be interpreted as any refutation of the hypothesis. Further research should explicitly measure health literacy and access to healthcare when investigating the link of education and the risk of PD.

An alternative explanation of the observed positive associations could be the negative associations of educational level with smoking and of smoking with risk of PD^7^. However, our subgroup analysis testing this did not explain much of the heterogeneity in effect sizes either.

Alternatively, the numerous significant results reported despite an effect close to unity in our meta-analysis might be explained statistically. Only 2 of the 24 studies included in the meta-analysis controlled for multiple comparisons although all studies investigated multiple risk factors. Moreover, although our meta-analysis does not seem to have been affected by small study effects, bias might still have distorted the image painted by some of the reports. Namely, many reports tested the effect of education on the risk of PD in various ways of which some were significant and others were not. Yet, in discussing their results, the reports highlighted the significant results and disregarded the insignificant tests. Lastly, the high heterogeneity in the operationalization of education might have additionally contributed to the heterogeneity in results. Henceforth, a combination of operationalization heterogeneity, type-I error inflation and selective emphasis might be the best explanation for the purported association of education and the risk of PD. Taken all together, this systematic review and meta-analysis does not support the hypothesis that higher educational attainment is associated with an increased risk of Parkinson’s disease.

## Data availability statement

All data used for the meta-analysis is available at https://osf.io/j9hp2. The script available at https://github.com/LaurenzLammer/pd_education_ma automatically loads this data.

## Authors roles

LL: Conceptualization, Data Curation, Formal Analysis, Funding acquisition, Investigation, Methodology, Software, Visualization, Writing – Original Draft Preparation, Writing – Review & Editing

CPP: Conceptualization, Data Curation, Methodology, Writing – Review & Editing

MK: Data Curation, Validation, Writing – Review & Editing

AS: Data Curation, Writing – Review & Editing

SB: Conceptualization, Data Curation, Methodology, Supervision, Writing – Review & Editing

## Appendix

### Database searches

#### PubMed

The search was performed on the 6^th^ of March 2025.

("Educational Status"[Mesh:NoExp] OR "Education*"[tiab] OR "Academic*" [tiab] OR "Social Class"[Mesh] OR "social class*"[tiab] OR "Socioeconomic Status"[tiab] OR "Socioeconomic Level*"[tiab] OR "Social gradient*"[tiab] OR "Epidemiologic Factors"[Mesh:NoExp] OR "Epidemiologic* Factor*"[tiab] OR "Determinant*"[tiab] OR "behavio* risk factor*"[tiab] OR "environmental risk factor*"[tiab] OR "Social Determinants of Health/statistics and numerical data"[Mesh])

AND ("Parkinson Disease"[Mesh] OR "Parkinson*" [tiab])

NOT ("Animals"[Mesh] NOT "Humans"[Mesh])

NOT ("Review"[pt] OR "Meta-Analysis" [pt] OR "Case Reports" [pt] OR "Clinical Study"[pt] OR "Comment"[pt] OR "Editorial"[pt] OR "Consensus Development Conference"[pt] OR "Guideline"[pt] OR "Evaluation Study"[pt])

#### PsychInfo

The search was performed on the 6^th^ of March 2025.

((Education/ or Academic Achievement/ or Education*.ti,ab. or exp Socioeconomic Status/ or social class*.ti,ab. or Socioeconomic Status.ti,ab. or Socioeconomic

Level*.ti,ab. or Social gradient*.ti,ab. or Epidemiology/ or Epidemiologic* Factor*.ti,ab. or behavio* risk factor*.ti,ab. or environmental risk factor*.ti,ab. or Social Determinants of Health/) and (exp "Parkinsons Disease"/ or Parkinson*.ti,ab.))

not (exp animal*/ or exp invertebrates/ or exp vertebrates/ or exp mice/ or exp "plant (botanical)"/)

not (exp Clinical Trial*/ or Meta Analysis/ or exp Literature Review/ or Treatment Guidelines/ or Case Report/ or exp Evaluation/)

not (Bibliography or "Column/Opinion" or "Comment/Reply" or Encyclopedia Entry or Interview or Editorial or Letter or Obituary or Poetry or Review-Book or Review-Media or Review-Software & Other or "Journal Column/Opinion" or Journal Column or Journal Editorial or Journal Editorials or Journal Information or Journal Letter or Journal Letters or Journal Obituaries or Journal Obituary or Journal Review-Book or Journal Review-Books or Journal Review-Other or "Journal Review-Software/Video/Other" or Journal Review-Software or Journal Review-Video).dt.

#### Web of Science

The search was performed on the 7^th^ of March 2025.

(TS=Education* OR TS=Academic* OR TS=Social Class* OR TS=Socioeconomic Status OR TS=Socioeconomic Level* OR TS=Social gradient* OR TS=Epidemiologic Factor* OR TS=Determinant* OR TS=behavio* risk factor* OR TS=environmental risk factor* OR TS=Social Determinants of Health)

AND (TS=Parkinson*)

NOT ((TS=Animal* OR TS=mice OR TS=Rats) NOT TS=Human*)

NOT ((TS=Child* OR TS=Infant OR TS=Adolescents) NOT TS=Adult)

NOT (TS=Review OR TS=Meta-Analysis OR TS=Case Report* OR TS=”case series” OR TS=”Clinical Study” OR TS=intervention OR TS=RCT OR TS=Comment OR TS=Editorial OR TS=Consensus OR TS=Guideline OR TS=”Evaluation study” OR TS=Patent OR TS=”Editorial Material” OR TS=”Clinical Trial” OR TS=Letter OR TS=Book OR TS=News OR TS=Biography OR TS=”Review Article” OR TS=Meeting OR TS=Correction OR TS=”Retracted Publication”)

NOT (LA=Spanish OR LA=French OR LA=Czech OR LA=Turkish OR LA=Portuguese OR LA=Hungarian OR LA=Italian OR LA=Polish OR LA=Russian OR LA=Korean OR LA=Persian OR LA=Croatian OR LA=Japanese) and Review Article or Editorial Material or Book Review or Retracted Publication or Data Paper or Music Performance Review

(Exclude – Document Types)

#### Embase

The search was performed on the 7^th^ of March 2025.

((exp education/ or exp educational status/ or ’education’.ab,kw,ti. or exp academic achievement/ or exp social class/ or ’social class’.ab,kw,ti. or exp socioeconomic status/ or ’socioeconomic status’.ab,kw,ti. or exp social determinants of health/ or ’social determinants of health’.ab,kw,ti. or ’behavioural risk factor’.ab,kw,ti. or ’behavioral risk factor’.ab,kw,ti. or ’epidemiological factor*’.ab,kw,ti. or exp social status/) and (exp Parkinson disease/ or ’Parkinson disease’.ab,kw,ti.))

not (exp animal/ not exp human/)

not (Conference review or Books or editorial or erratum or letter or note or Review or Short survey).pt.

limit 1 to (human and (english or german))

### Extracted information

We collected data on:

∘ Metadata: extractor, extraction date, extraction version number, title, author(s), date of publication, doi and/or other identifier(s)
∘ Participants: Total number of participants (number of cases and controls, at each timepoint, if applicable), setting, geographical location, study eligibility criteria, descriptive statistics
∘ Education: assessment method, variable type with cut-offs
∘ PD: Ascertainment method, assessor qualification, diagnostic criteria, symptom severity assessment method, timepoint of assessment
∘ Study design: study name, general study design, recruitment and sampling procedures, enrolment start and end dates, length of follow-up (if applicable), methods used to prevent and control for confounding, information biases, measures taken to control selection biases, preregistration, preventing/handling missing data
∘ Statistical analysis: unit of analysis, statistical methods, multiplicity control, a priori power calculation
∘ Results: summary data, between-group estimates with precision
∘ Other: funding, conflicts of interest, key conclusions of the study authors, need for correspondence, miscellaneous comments

### Country wise income categorization

Our categorizations were based on The World Bank’s World Development Indicators^104^. High income countries were dichotomously distinguished from all other categories. Due to the high missingness of information on and heterogeneity in recruitment times, we used the publication year to discern the appropriate category for each study’s population. A study with samples from numerous Latin American countries was summatively categorized in the low- and middle-income group and mendelian randomization studies of participants of European ancestry were grouped in the high-income category.

### Effect size homogenization

If education was treated as a continuous variable with the metric e.g. in years of schooling, we assumed the average difference in years of schooling between the low and the high education group to be five years. Accordingly, the OR per year of schooling was converted: OR_p.a._^5^ = OR_dichotomous_. Likewise, if education was coded in levels and effect sizes were provided per level the effect size was taken to the power of n (of levels) / 2. If there were 3 levels, this would mean OR_per level_^1.5^ = OR_dichotomous_.

^11,15,53,55–57,62,68,71,73^ provided ORs from multiple comparisons that were aggregated using the aggregate function of **{metafor}** setting ρ (the correlation of the sampling errors) to 0.6.

^59–61,64^ provided mendelian randomization based OR estimates derived from different methods. These estimates were also aggregated using the aggregate function of **{metafor}** setting ρ (the correlation of the sampling errors) to 0.6.

ORs for ^58,74^ were calculated from raw count data of cases and controls using the oddsratio() function of **{epitools}** employing the default median-unbiased estimation.

ORs and confidence intervals for ^64^ were calculated from beta coefficients and SEs from logistic regression using the following formulae: OR = e^beta^. Lower limit = e^beta – 1.96 * SE^. Upper limit = e^beta + 1.96 * SE^.

Standard errors for studies that reported ORs with p-values ^65,69^ were calculated using: SE = log(OR) / z, where z is the quantile from the standard normal distribution of 1 – p/2.

^17^ provided standardized incidence ratios for different education levels. Using either that for <9 years or that for >16 years would clearly indicate the direction of the effect. As the <9 years group is more than 10 times as large as the >16 years group we chose the former as our measure of effect size for this study. The separate effect estimates for men and women were aggregated as in the other studies.

### Linked reports

Both ^12^ and ^52^ are based on the Rochester Epidmiology project. For the meta-analysis we included Frigerio et al. as Savica et al.’s gender stratified analysis did not fit our research question as well.

Both ^16^ and ^65^ used data from the UK Biobank. As preregistered, we based our choice on the RoB as determined by the QUIPS tool. This led us to choose Jacobs et al.

^17,56,73^ are based on overlapping Swedish registries. As the samples are merely overlapping, we included all studies in our main analysis but conducted a sensitivity analysis only including the Swedish register-based study with the lowest RoB (Li et al. 2009).

Furthermore, three mendelian randomization studies^59,60,64^ were based on GWAS from the same studies (education is based on:^105^; PD is based on: ^106^). Of these studies we chose Li et al. based on its RoB. Additionally, ^61^ also relied on ^106^ for the PD GWAS but used ^107^ for the education GWAS and was hence included in the MA.

Moreover, both ^82^ and ^78^ are based on the Yonsei Parkinson Center database. As both studies only concern our secondary outcome, that is not analysed meta-analytically, this was discussed in the narrative summary.

All studies (except for Savica et al. 2013 because of bad fit with research question) are maintained in the published dataset so that other researchers can explore the impact of including/excluding individual studies.

### Reflexivity

A few years ago, while reading Medicine, I (LL) worked in a neurological ward during semester break. Studying the list of our in-patients, I was surprised to find that almost all PD patients had a “Dr.” in front of their names. I was surprised again when I read that multiple studies found a positive association of education with risk of PD in larger samples than our little ward. This was and is very counterintuitive to me as I struggle to find an explanation for how more education, with all the privileges it entails, may lead to a greater risk of PD. As a social epidemiologist, I was, at the outset of this research project, unconvinced by an actual adverse effect of more education and held a privileged access to healthcare to be the more plausible explanation of the association. Beyond limited clinical experience with PD, I have neither first-hand experience of PD or a close friend or relative suffering from the ailment.

## Supplementary Figures

**Supplementary Figure 1:**
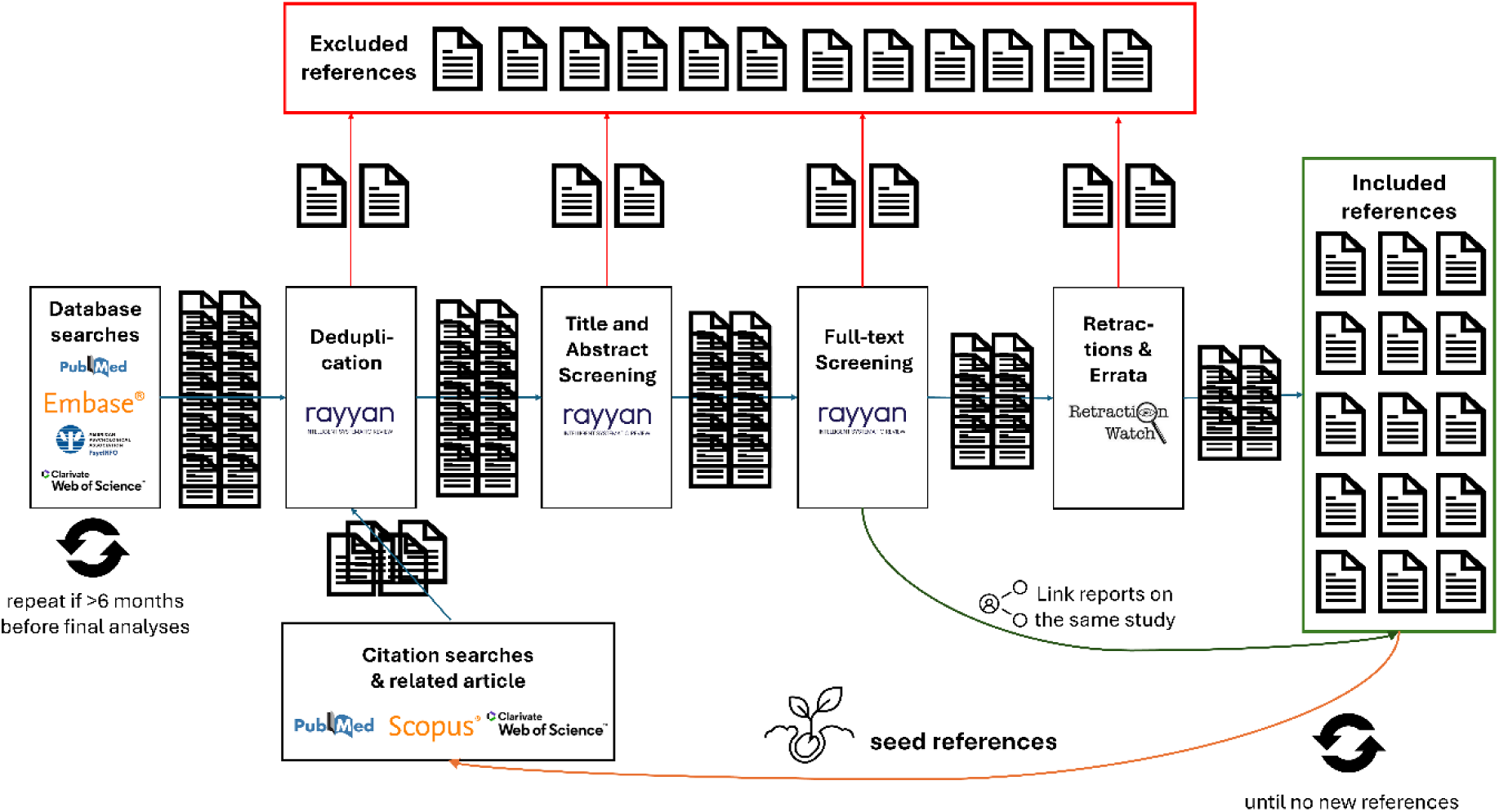
Overview of the search and screening strategies.

**Supplementary Figure 2:**
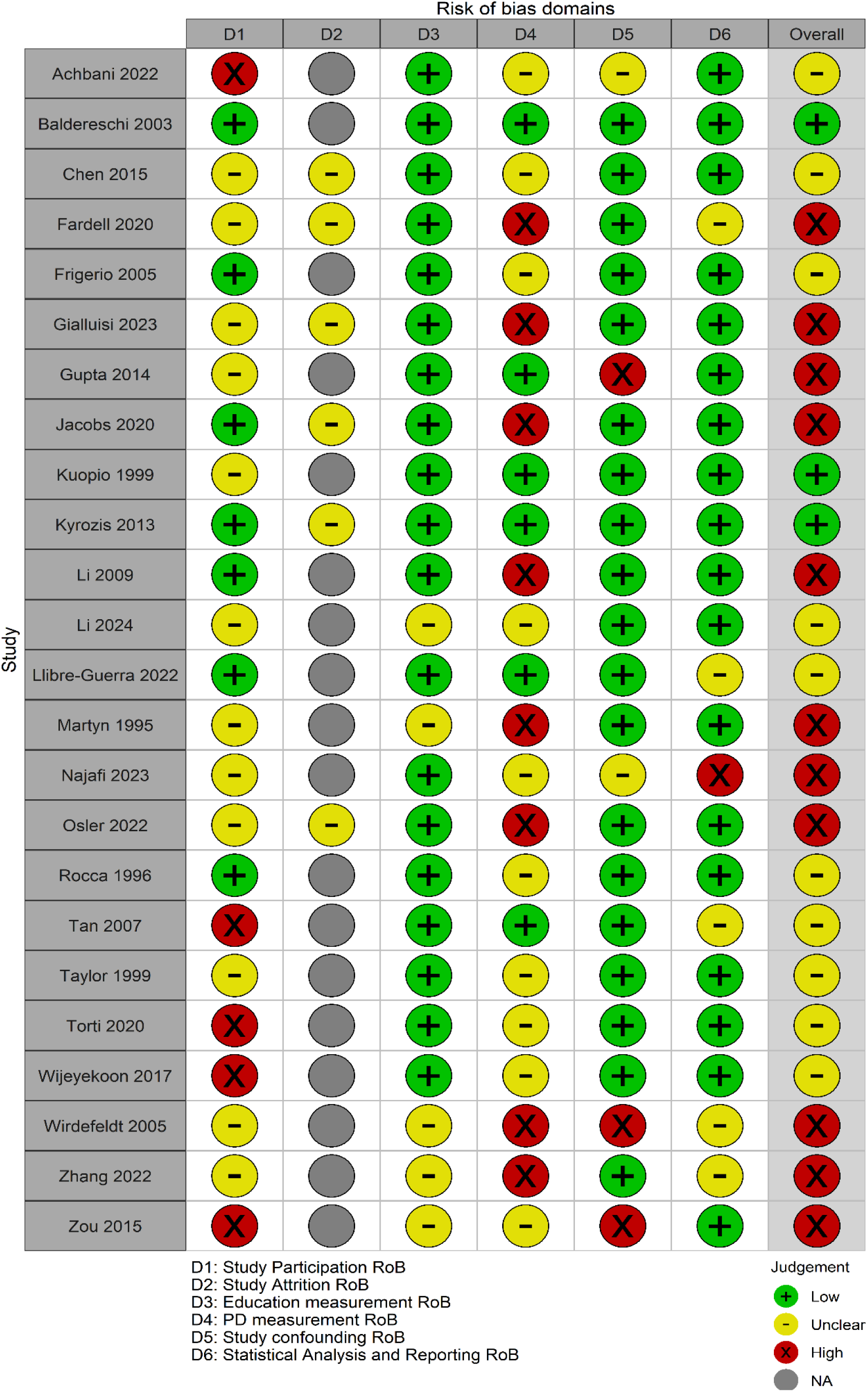
Traffic light plot of the domain-wise and overall RoB of each study included in our main meta-analysis.

**Supplementary Figure 3:**
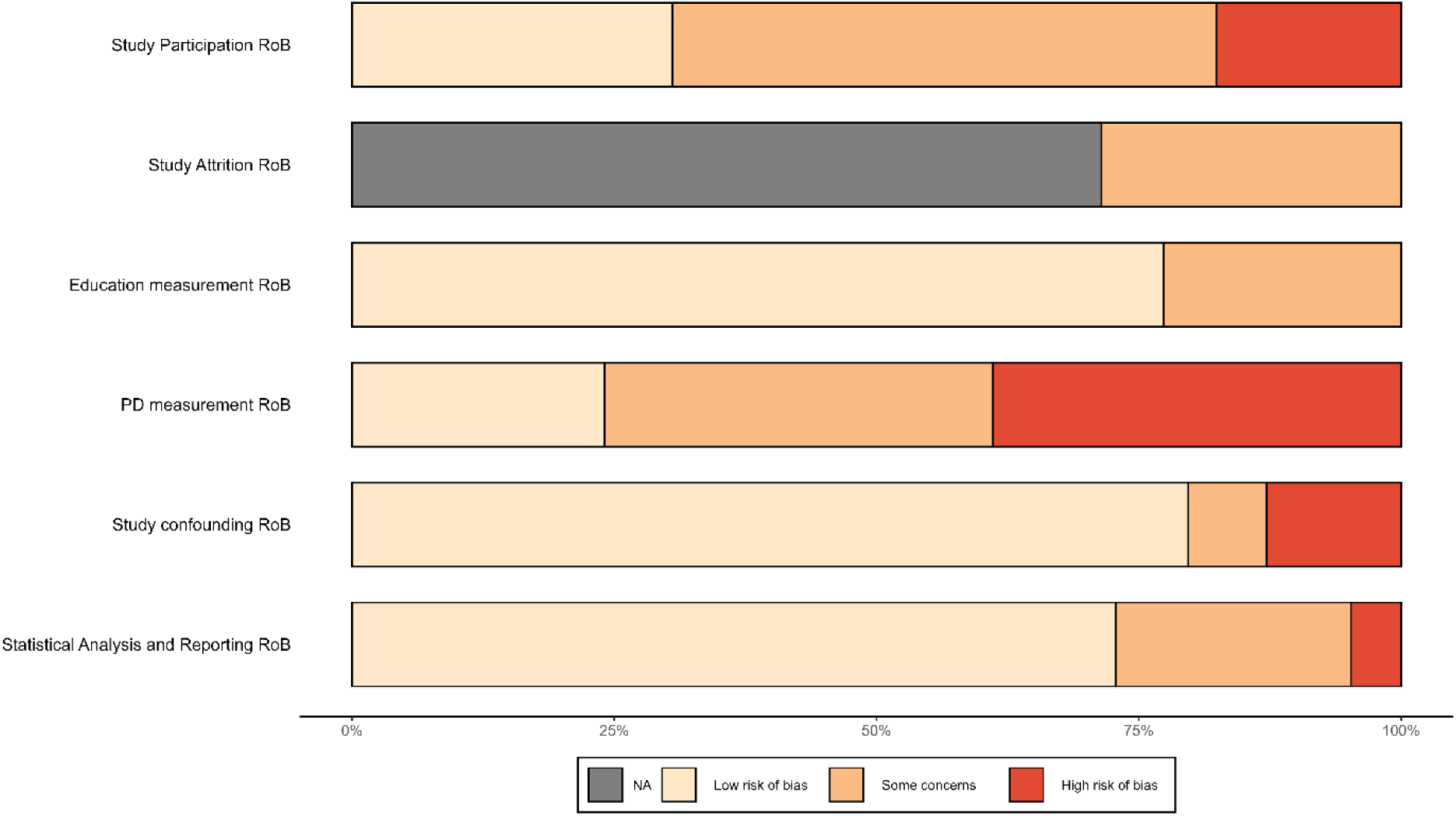
Weighted domain-wise RoB of our main analysis.

**Supplementary Figure 4:**
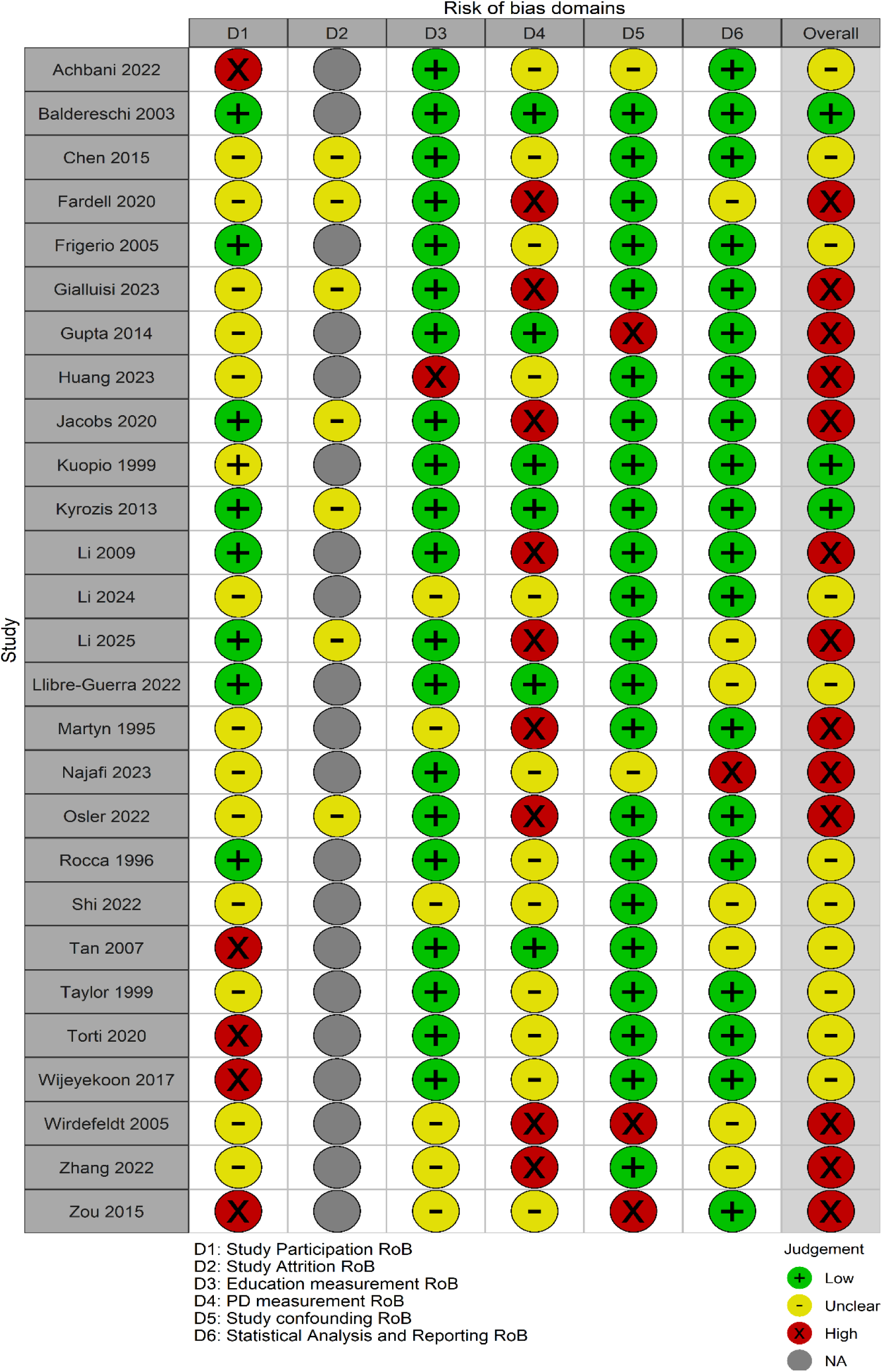
Traffic light plot of domain-wise and overall RoB of all reports relevant to our primary outcome including linked reports.

**Supplementary Figure 5:**
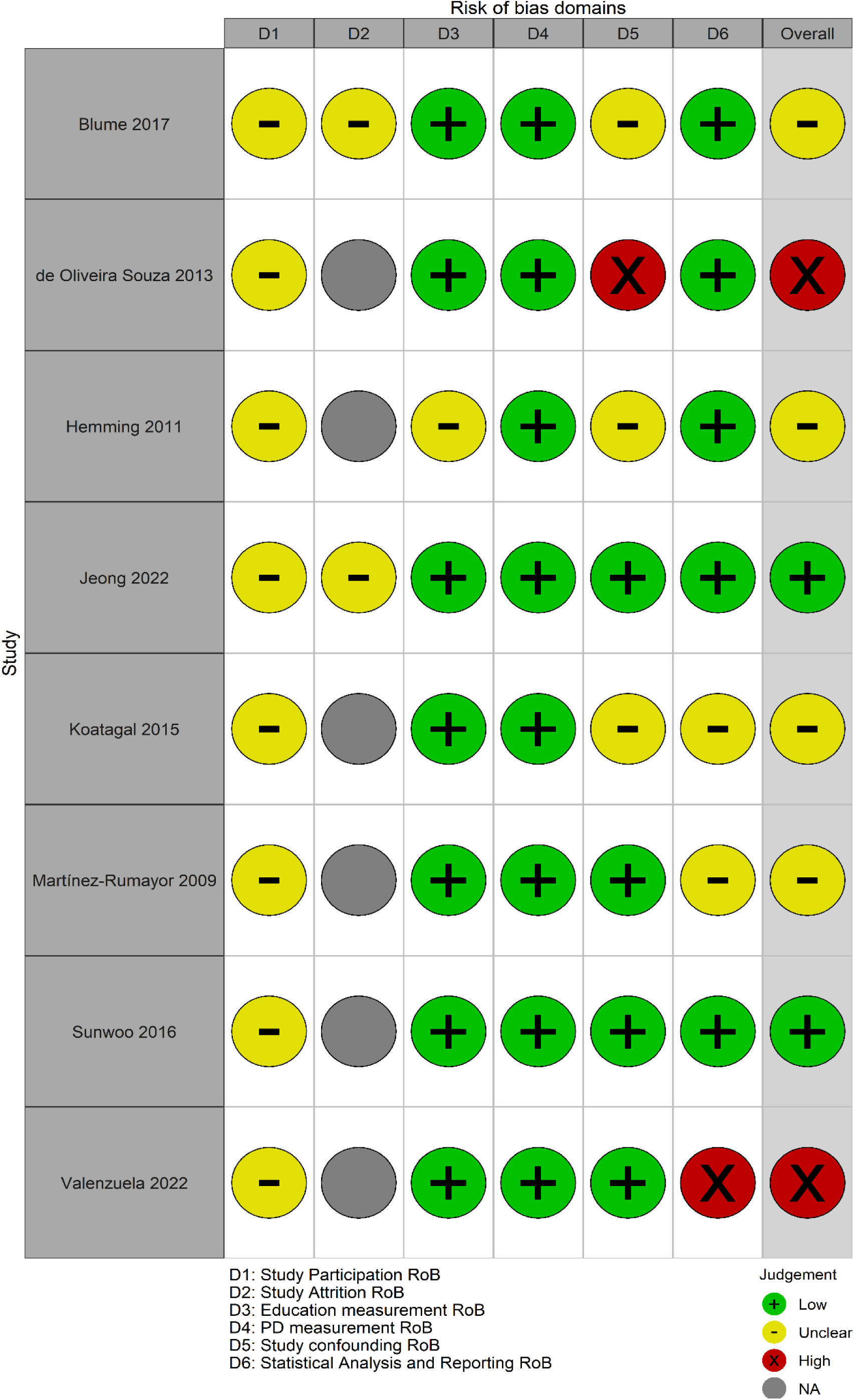
Traffic light plot of studies relevant to our secondary outcome.

**Supplementary Figure 6:**
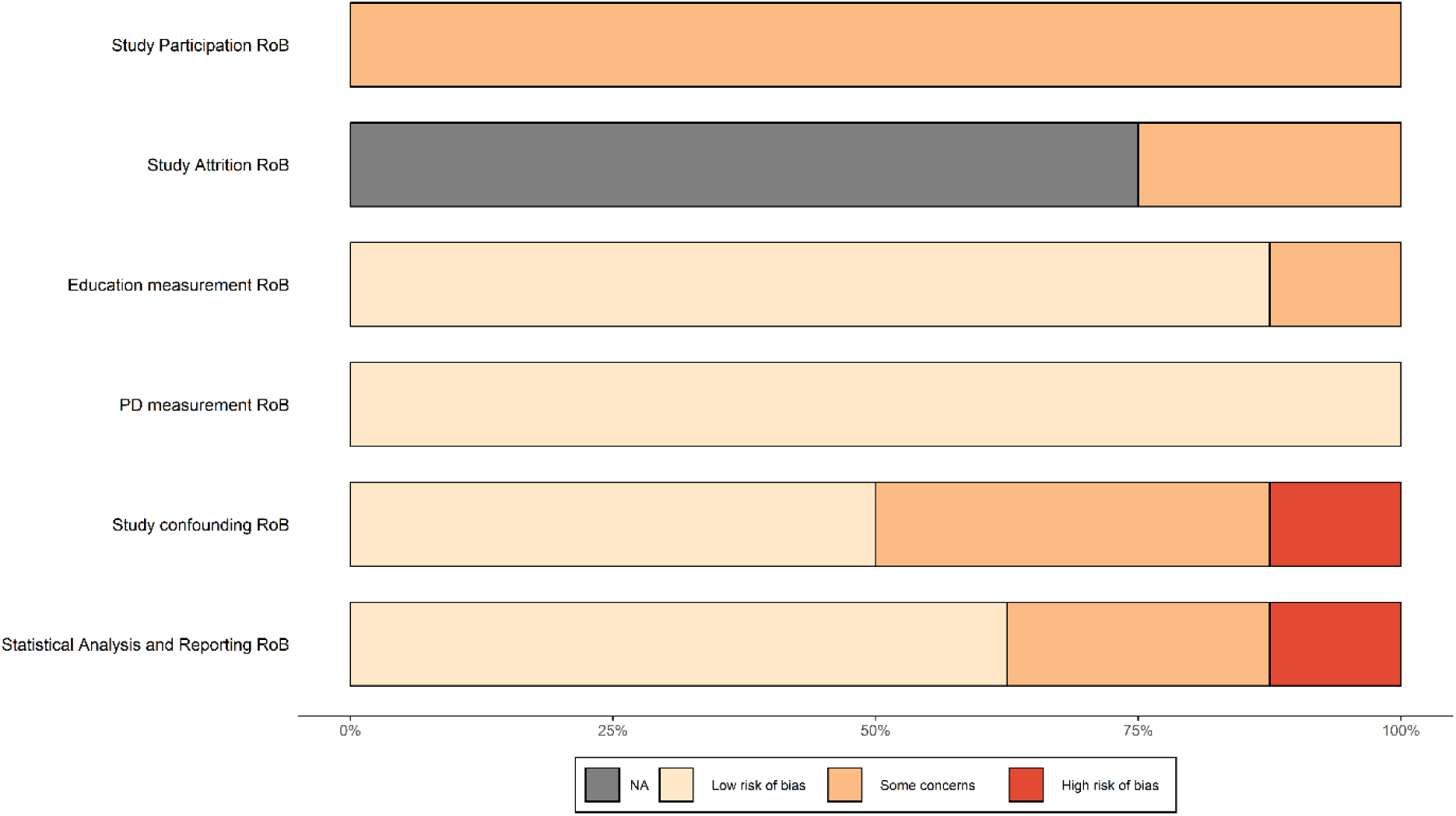
Unweighted domain-wise RoB for our secondary outcome.

**Supplementary Figure 7:**
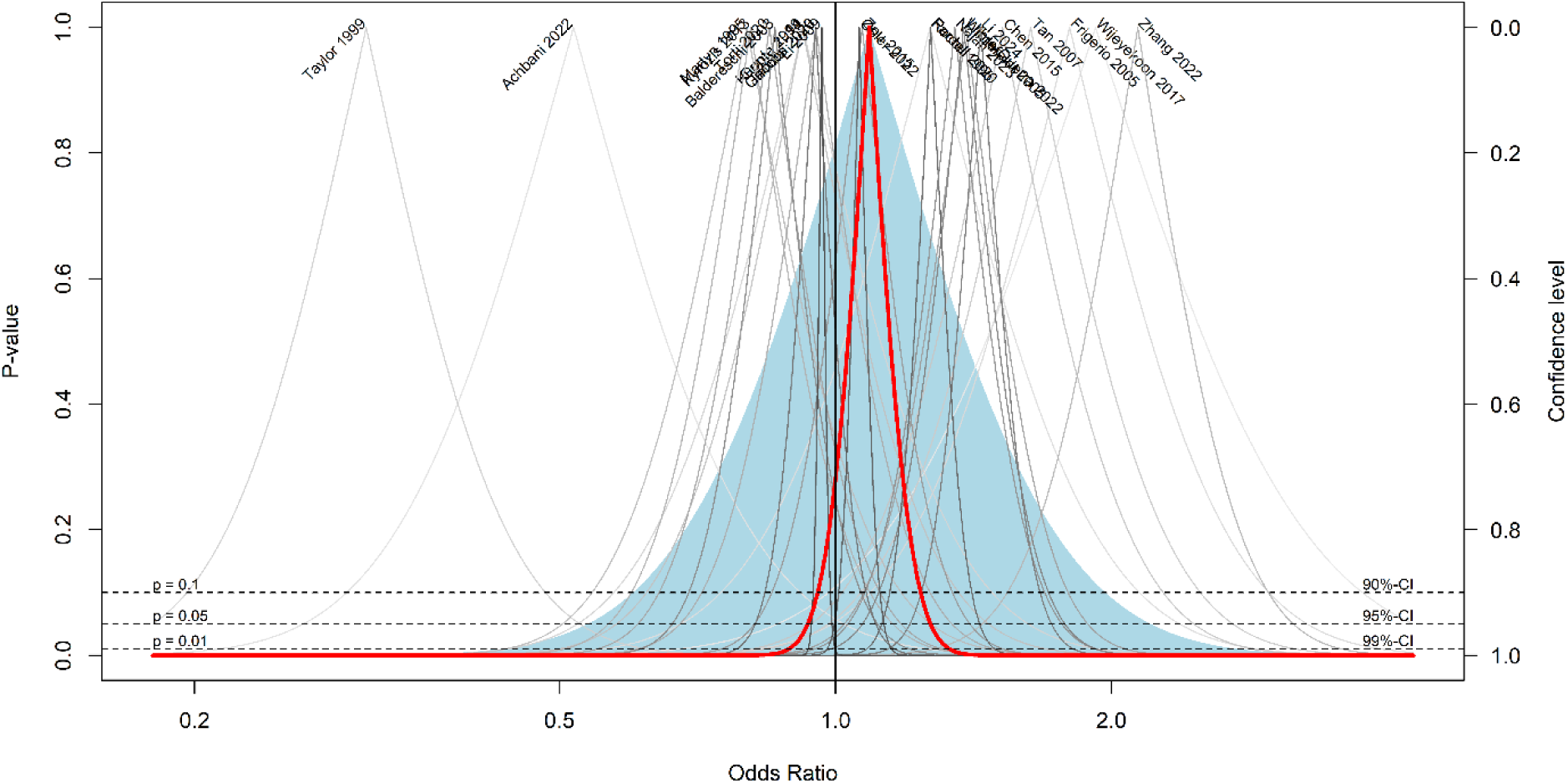
Drapery plot of our main meta-analysis. The dashed lines indicate different levels of significance. The thin grey lines indicate the effects of individual studies and the thicker red line represents the overall effect.

**Supplementary Figure 8:**
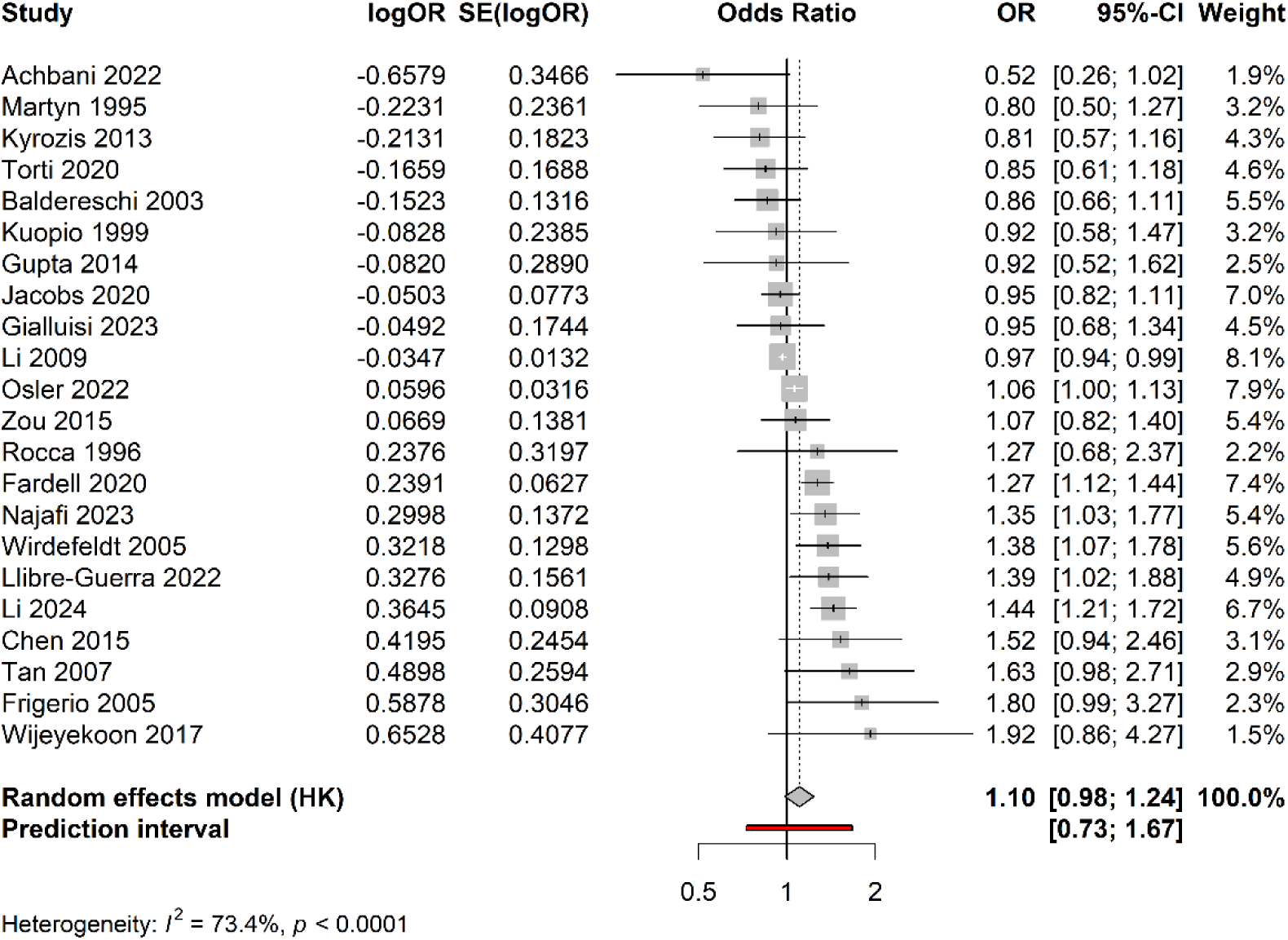
Forest plot of the meta-analysis excluding outliers.

**Supplementary Figure 9:**
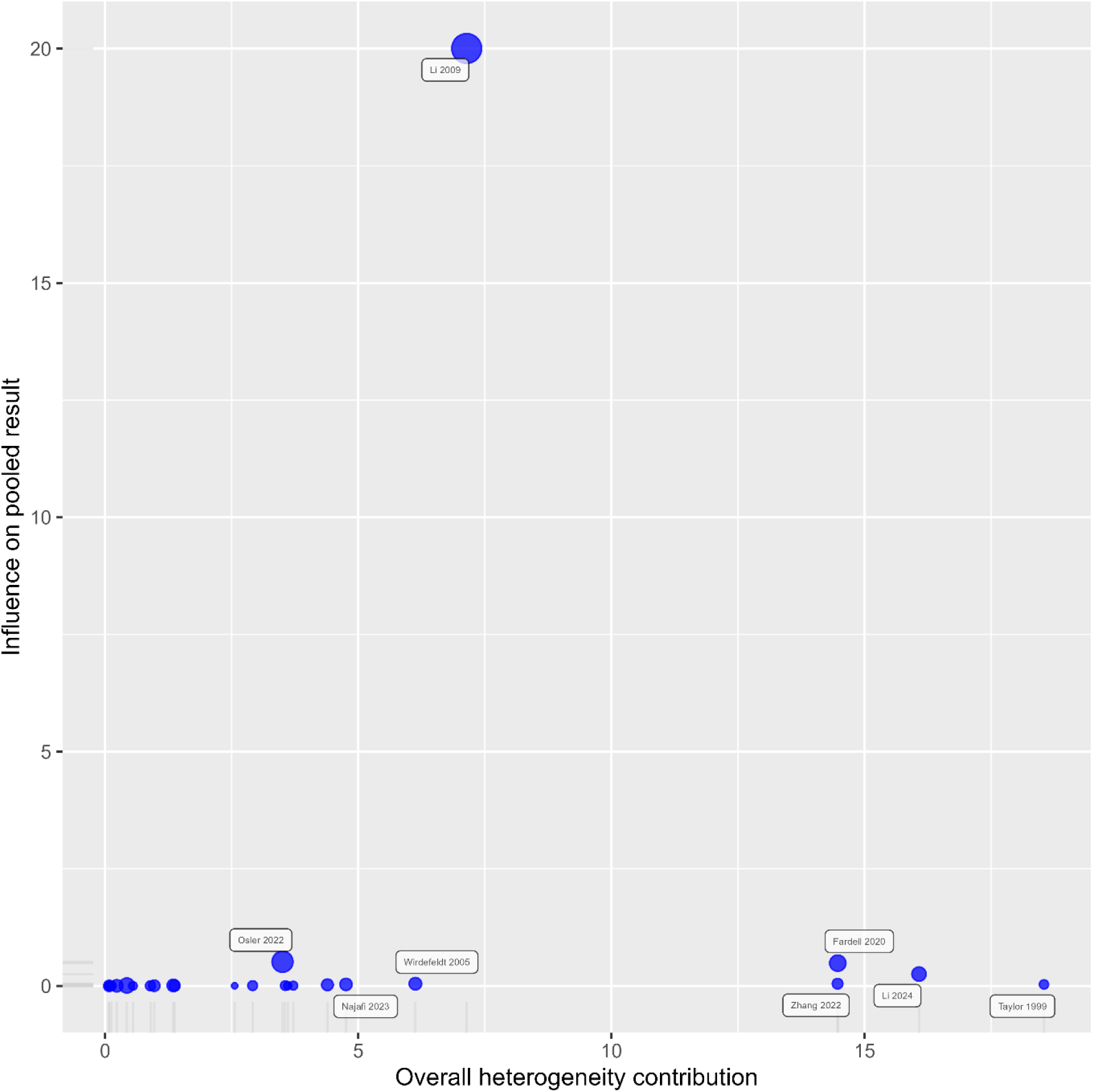
Baujat plot of the studies included in our main meta-analysis. The circle diameter corresponds to the studies relative weight.

**Supplementary Figure 10:**
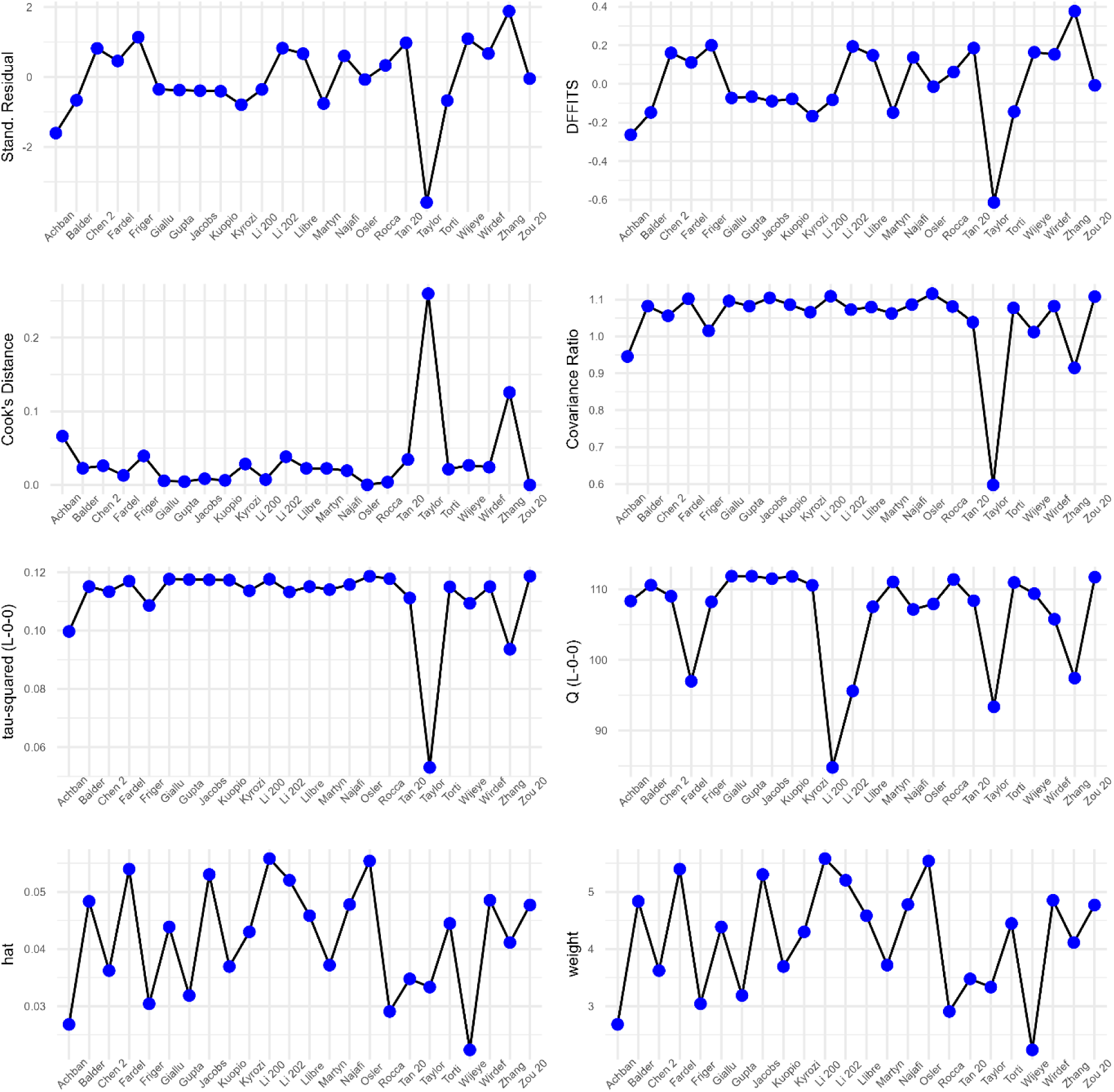
Depictions of multiple measures of individual study influence. Influential cases would be depicted in red but there are none.

**Supplementary Figure 11:**
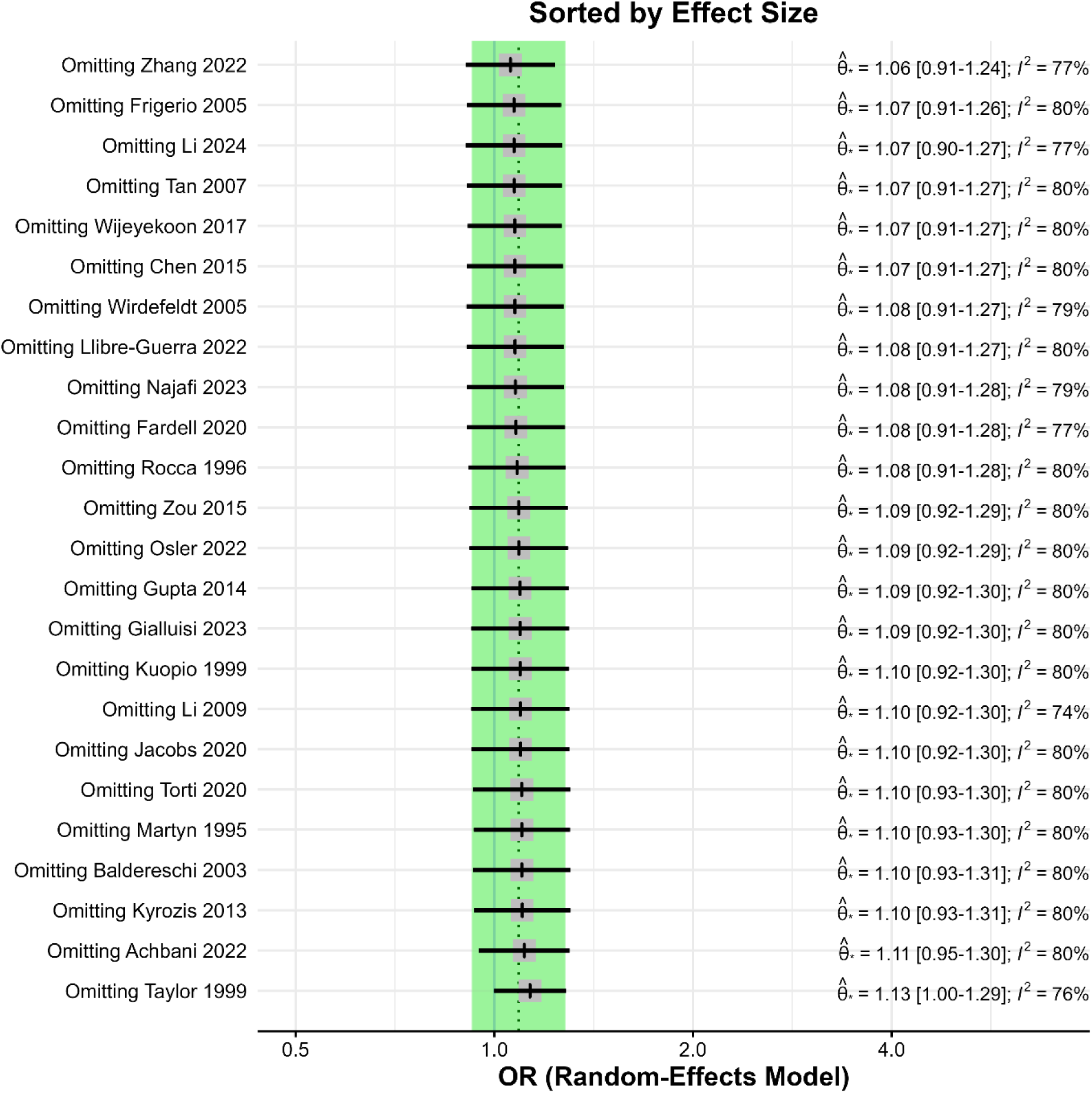
Results of leave-one-out meta-analyses sorted by effect size.

**Supplementary Figure 12:**
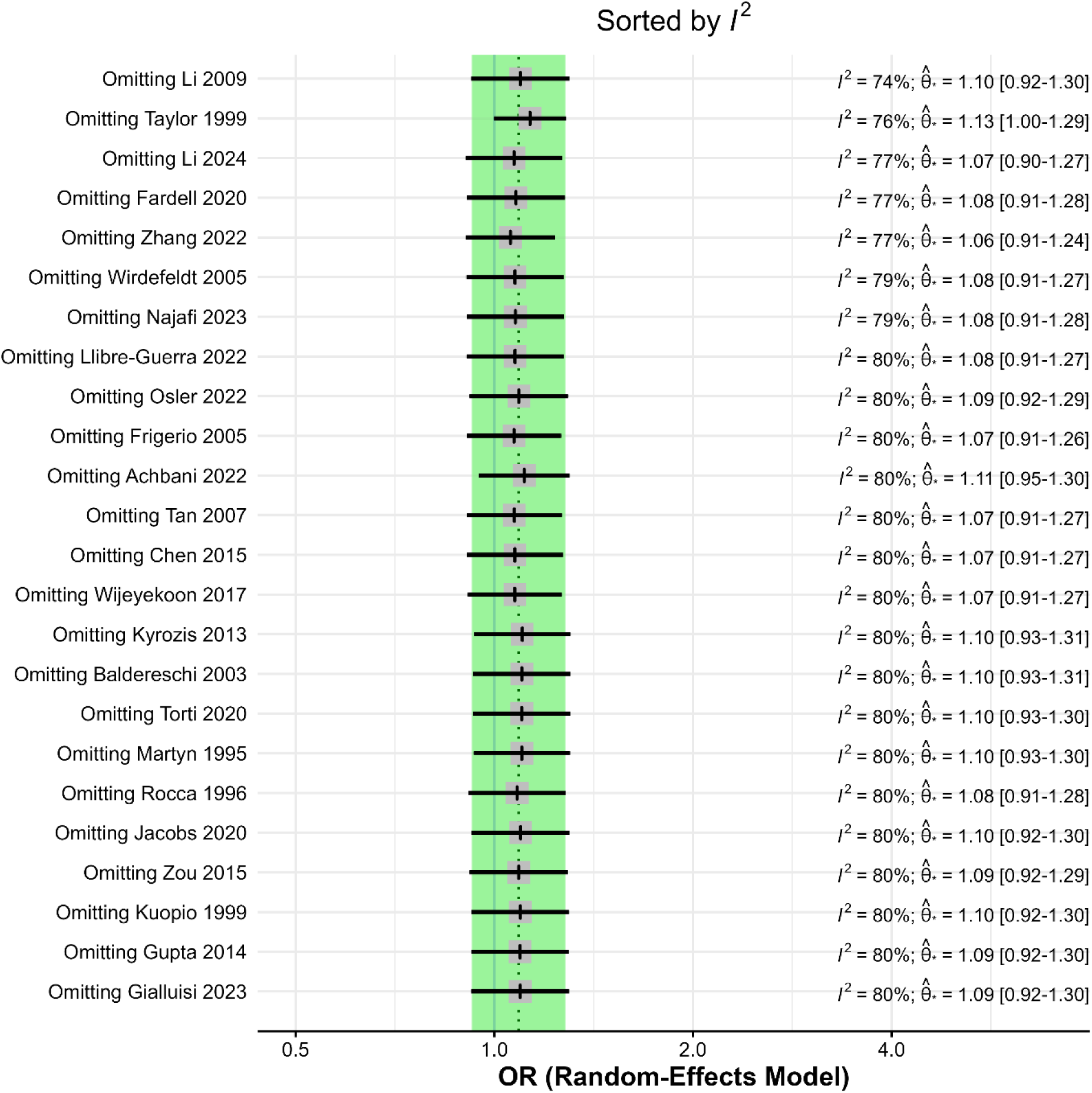
Results of leave-one-out meta-analyses sorted by I^2^.

**Supplementary Figure 13:**
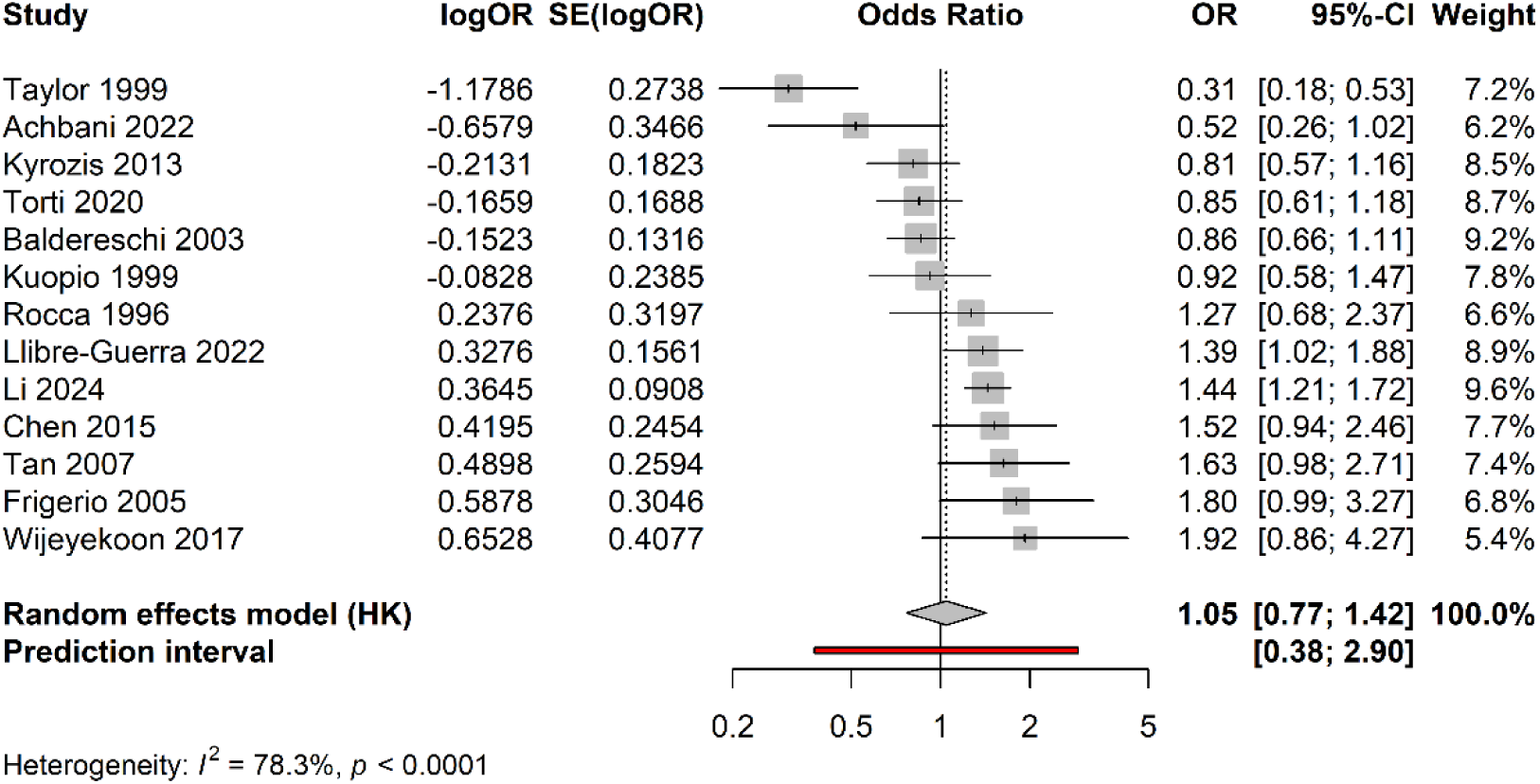
Forest plot of meta-analysis excluding studies at high RoB.

**Supplementary Figure 14:**
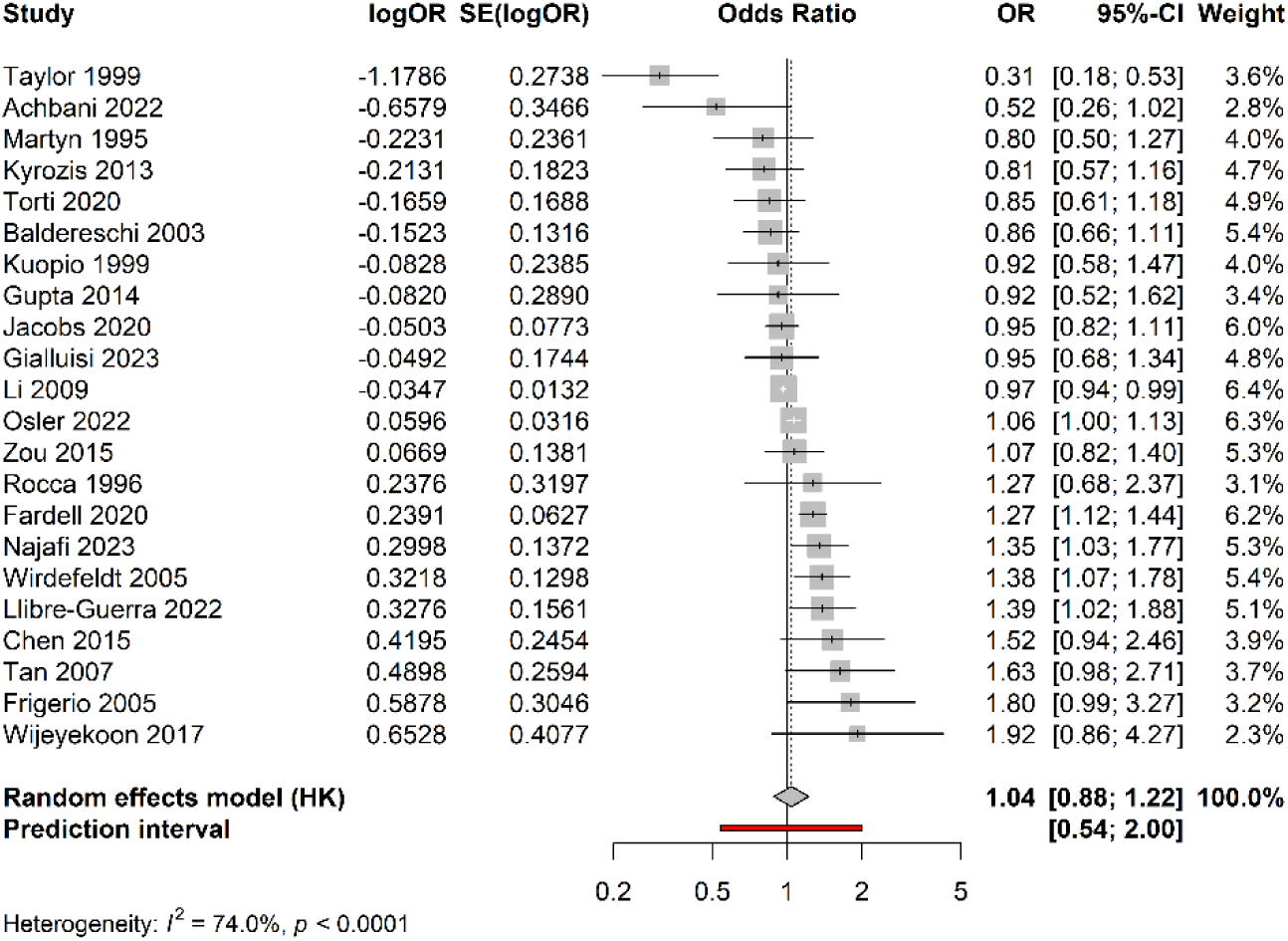
Forest plot of meta-analysis excluding mendelian randomization studies.

**Supplementary Figure 15:**
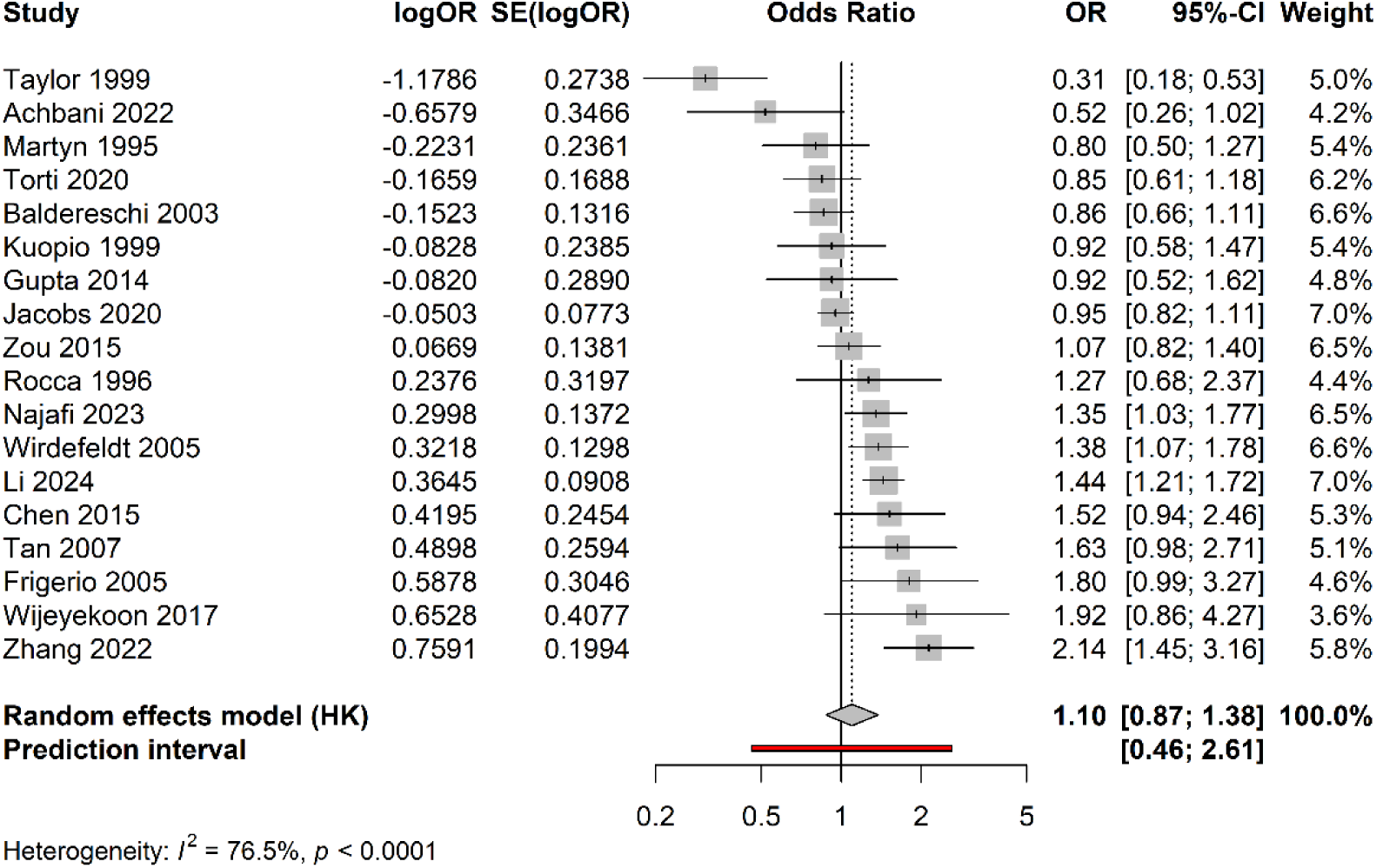
Forest plot of meta-analysis excluding studies reporting effect measures other than OR.

**Supplementary Figure 16:**
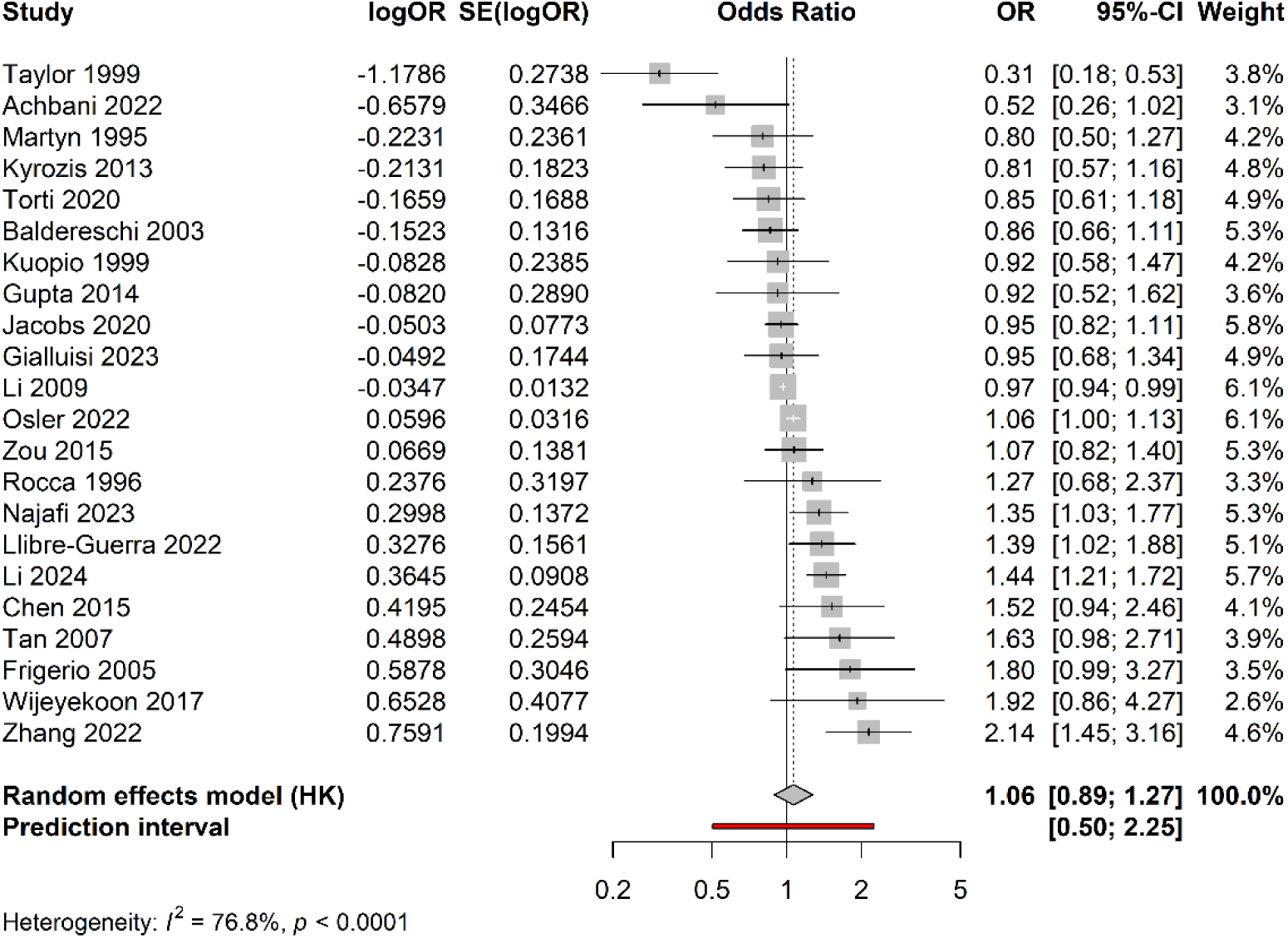
Forest plot of meta-analysis including only the Swedish register-based study with the lowest RoB.

**Supplementary Figure 17:**
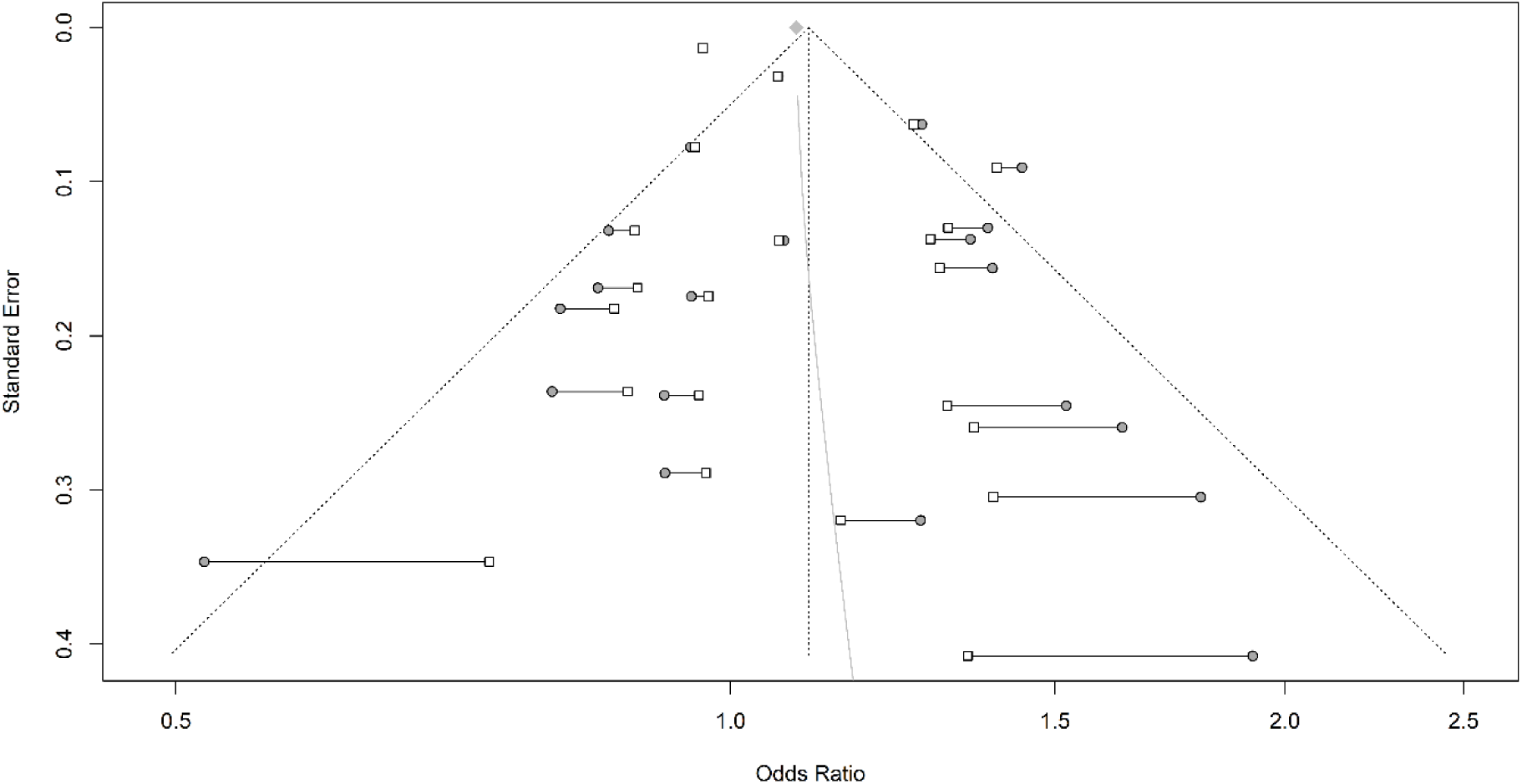
Funnel plot of the limit meta-analysis excluding outlying studies. Circles depict original effect sizes and squares the effect sizes after adjustment. The light grey line indicates the effect size at different standard errors.

**Supplementary Figure 18:**
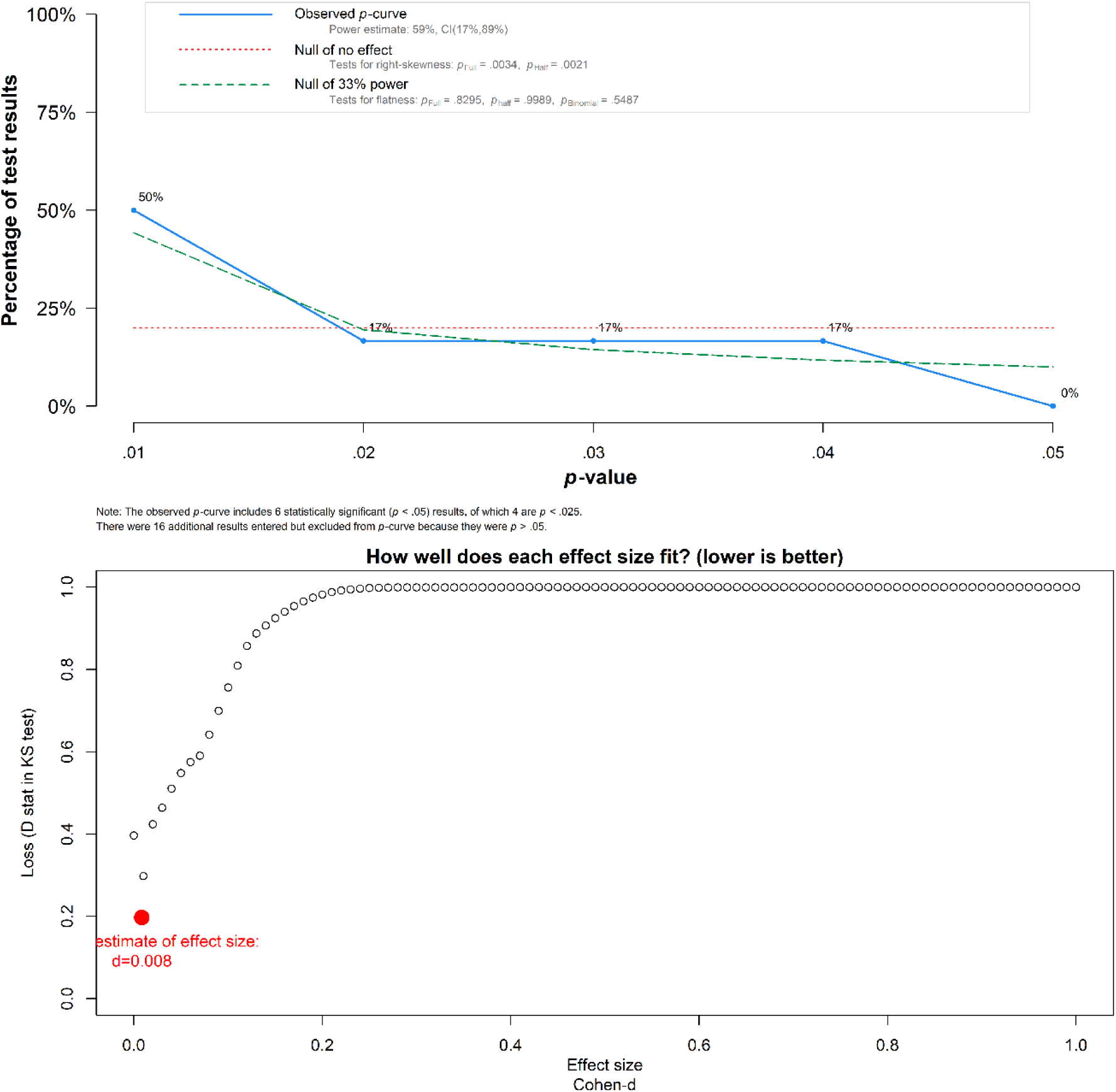
The upper figure shows the standard p-curve (blue) compared to a null effect (red, dotted) and a true effect at insufficient power (green, dashed). The lower figure shows the adjusted effect size estimate. The latter is unreliable due to the high heterogeneity, though.

## Supplementary tables

**Supplementary Table 1.**

| Authors | Year | Reason for exclusion |
| --- | --- | --- |
| Dick et al. | 2007 | Education wasn't a predictor of interest |
| Torrealba-Acosta et al. | 2021 | Education wasn't a predictor of interest |
| Georgiou et al. | 2019 | Education wasn't a predictor of interest |
| Marder et al. | 1998 | Education wasn't a predictor of interest |
| Nicoletti et al. | 2010 | Wrong type of study |
| Belvisi et al. | 2020 | Education wasn't a predictor of interest |
| Alcalay et al. | 2011 | Education wasn't a predictor of interest |
| Sohn et al. | 2016 | Wrong publication type |
| Yang et al. | 2014 | Investigated SES not education |
| Zhu et al. | 2024 | Wrong publication type |

**Supplementary Table 2.** Characteristics of the included studies for the secondary outcome.

| **Characteristics of the included studies for the secondary outcome** |  |  |  |
| --- | --- | --- | --- |
| **Report** | **Location** | **Sample size** | **effect metric** |
| Blume 2017 | Germany | 102 | UPDRS III |
| de Oliveira Souza 2013 | Brazil | 58 | H & Y, UPDRS I & III, BBS |
| Hemming 2011 | USA | 1,159 | UPDRS total, UPDRS III |
| Jeong 2022 | South Korea | 478 | UPDRS III, LID HR |
| Kotagal 2015 | USA | 142 | UPDRS III |
| Martínez-Rumayor 2009 | Mexico | 247 | age at symptom onset |
| Sunwoo 2016 | South Korea | 182 | UPDRS III |
| Valenzuela 2022 | Spain | 40 | Gait speed |

UPDRS III = Unified Parkinson's Disease Rating Scale motor subscale; H & Y = Hoehn & Yahr stage; BBS = Berg Balance Scale; LID HR = Levodopa-induced dyskinesia hazard ratio;

**Supplementary Table 3.** Risk-of-bias judgment for each of six domains of bias, and for the overall risk of bias of studies on our primary outcome.

|  |  |  |  |  |  |  |  |
| --- | --- | --- | --- | --- | --- | --- | --- |
| report | Study Participation | Study Attrition | Education Measurement | PD Measurement | Confounding | Statistical Analysis and Reporting | Overall RoB |
| Achbani 2022 | high RoB |  | low RoB | moderate RoB | moderate RoB | low RoB | moderate RoB |
| Baldereschi 2003 | low RoB |  | low RoB | low RoB | low RoB | low RoB | low RoB |
| Chen 2015 | moderate RoB | moderate RoB | low RoB | moderate RoB | low RoB | low RoB | moderate RoB |
| Fardell 2020 | moderate RoB | moderate RoB | low RoB | high RoB | low RoB | moderate RoB | high RoB |
| Frigerio 2005 | low RoB |  | low RoB | moderate RoB | low RoB | low RoB | moderate RoB |
| Gialluisi 2023 | moderate RoB | moderate RoB | low RoB | high RoB | low RoB | low RoB | high RoB |
| Gupta 2014 | moderate RoB |  | low RoB | low RoB | high RoB | low RoB | high RoB |
| Huang 2023 | moderate RoB |  | high RoB | moderate RoB | low RoB | low RoB | high RoB |
| Jacobs 2020 | low RoB | moderate RoB | low RoB | high RoB | low RoB | low RoB | high RoB |
| Kuopio 1999 | low RoB |  | low RoB | low RoB | low RoB | low RoB | low RoB |
| Kyrozis 2013 | low RoB | moderate RoB | low RoB | low RoB | low RoB | low RoB | low RoB |
| Li 2009 | low RoB |  | low RoB | high RoB | low RoB | low RoB | high RoB |
| Li 2024 | moderate RoB |  | moderate RoB | moderate RoB | low RoB | low RoB | moderate RoB |
| Li 2025 | low RoB | moderate RoB | low RoB | high RoB | low RoB | moderate RoB | high RoB |
| Llibre-Guerra 2022 | low RoB |  | low RoB | low RoB | low RoB | moderate RoB | moderate RoB |
| Martyn 1995 | moderate RoB |  | moderate RoB | high RoB | low RoB | low RoB | high RoB |
| Najafi 2023 | moderate RoB |  | low RoB | moderate RoB | moderate RoB | high RoB | high RoB |
| Osler 2022 | moderate RoB | moderate RoB | low RoB | high RoB | low RoB | low RoB | high RoB |
| Rocca 1996 | low RoB |  | low RoB | moderate RoB | low RoB | low RoB | moderate RoB |
| Shi 2022 | moderate RoB |  | moderate RoB | moderate RoB | low RoB | moderate RoB | moderate RoB |
| Tan 2007 | high RoB |  | low RoB | low RoB | low RoB | moderate RoB | moderate RoB |
| Taylor 1999 | moderate RoB |  | low RoB | moderate RoB | low RoB | low RoB | moderate RoB |
| Torti 2020 | high RoB |  | low RoB | moderate RoB | low RoB | low RoB | moderate RoB |
| Wijeyekoon 2017 | high RoB |  | low RoB | moderate RoB | low RoB | low RoB | moderate RoB |
| Wirdefeldt 2005 | moderate RoB |  | moderate RoB | high RoB | high RoB | moderate RoB | high RoB |
| Zhang 2022 | moderate RoB |  | moderate RoB | high RoB | low RoB | moderate RoB | high RoB |
| Zou 2015 | high RoB |  | moderate RoB | moderate RoB | high RoB | low RoB | high RoB |

RoB = risk of bias; PD = parkinson's disease
Because they reported data from the same study as other reports, Fardell 2020, Huang 2023, Li 2024, Li 2025, Wirdefeldt 2005 were not included in the meta-analysis.

**Supplementary Table 4.** Risk-of-bias judgment for each of six domains of bias, and for the overall risk of bias of studies on our secondary.

| Risk-of-bias judgment for each of six domains of bias, and for the overall risk of bias of studies on our secondary outcome |  |  |  |  |  |  |  |
| --- | --- | --- | --- | --- | --- | --- | --- |
| report | Study Participation | Study Attrition | Education Measurement | PD Measurement | Confounding | Statistical Analysis and Reporting | Overall RoB |
| Blume 2017 | moderate RoB | moderate RoB | low RoB | low RoB | moderate RoB | low RoB | moderate RoB |
| de Oliveira Souza 2013 | moderate RoB |  | low RoB | low RoB | high RoB | low RoB | high RoB |
| Hemming 2011 | moderate RoB |  | moderate RoB | low RoB | moderate RoB | low RoB | moderate RoB |
| Jeong 2022 | moderate RoB | moderate RoB | low RoB | low RoB | low RoB | low RoB | low RoB |
| Koatagal 2015 | moderate RoB |  | low RoB | low RoB | moderate RoB | moderate RoB | moderate RoB |
| Martínez-Rumayor 2009 | moderate RoB |  | low RoB | low RoB | low RoB | moderate RoB | moderate RoB |
| Sunwoo 2016 | moderate RoB |  | low RoB | low RoB | low RoB | low RoB | low RoB |
| Valenzuela 2022 | moderate RoB |  | low RoB | low RoB | low RoB | high RoB | high RoB |

RoB = risk of bias; PD = parkinson's disease
Sunwoo 2016 and Jeong 2022 are based on the same registry.

**Supplementary Table 5.** Results of meta analyses excluding outliers.

| Results of meta-analyses excluding outliers |  |  |  |  |  |  |  |  |  |  |
| --- | --- | --- | --- | --- | --- | --- | --- | --- | --- | --- |
| Analysis | n studies | OR (95% CI) | t-/z-value | p-value | prediction interval | tau<sup>2</sup> | I<sup>2</sup> (%) | Q / Q'-Q | df | p-value of Q |
| main | 22 | 1.1 (0.98 - 1.24) | 1.784 | 0.089 | 0.73 - 1.67 | 0.037 | 73.4 | 78.919 | 21 | 0.000 |
| subgroup smoking | 22 | 1.1 (0.98 - 1.24) | 1.784 | 0.089 | 0.73 - 1.67 | 0.041 | 72.1 | 0.001 | 1 | 0.973 |
| smoking not controlled | 16 | 1.1 (0.97 - 1.26) | 1.607 | 0.129 | 0.7 - 1.74 | 0.041 | 70.1 | 50.137 | 20 | 0.000 |
| smoking controlled | 6 | 1.1 (0.79 - 1.53) | 0.737 | 0.494 | 0.61 - 1.98 | 0.041 | 76.9 | 21.602 |  | 0.001 |
| subgroup representative | 22 | 1.1 (0.98 - 1.24) | 1.784 | 0.089 | 0.73 - 1.67 | 0.036 | 62.3 | 1.036 | 1 | 0.309 |
| not representative | 9 | 1.02 (0.8 - 1.29) | 0.142 | 0.890 | 0.62 - 1.66 | 0.036 | 54.0 | 17.378 | 20 | 0.026 |
| representative | 13 | 1.15 (1.01 - 1.31) | 2.273 | 0.042 | 0.74 - 1.78 | 0.036 | 66.4 | 35.707 |  | 0.000 |
| subgroup PD assessment | 22 | 1.1 (0.98 - 1.24) | 1.784 | 0.089 | 0.73 - 1.67 | 0.037 | 72.8 | 0.977 | 1 | 0.323 |
| not all assessed | 14 | 1.07 (0.92 - 1.23) | 0.956 | 0.357 | 0.69 - 1.65 | 0.037 | 78.4 | 60.208 | 20 | 0.000 |
| all assessed | 8 | 1.2 (0.95 - 1.5) | 1.877 | 0.103 | 0.71 - 2 | 0.037 | 47.0 | 13.205 |  | 0.067 |
| subgroup income | 22 | 1.1 (0.98 - 1.24) | 1.784 | 0.089 | 0.73 - 1.67 | 0.034 | 71.0 | 1.483 | 1 | 0.223 |
| HIC | 8 | 1.23 (0.94 - 1.62) | 1.856 | 0.106 | 0.75 - 2.04 | 0.034 | 43.6 | 12.420 | 20 | 0.088 |
| L/MIC | 14 | 1.06 (0.93 - 1.2) | 0.938 | 0.365 | 0.69 - 1.61 | 0.034 | 77.0 | 56.513 |  | 0.000 |
| limit meta-analysis | 22 | 1.09 (0.93 - 0.93) | 1.054 | 0.292 |  | 0.037 | 73.4 | 13.900 | 1 | 0.000 |

OR (95% CI): Odds ratio with 95% confidence interval (adjusted for limit meta-analysis row); t/z-value: z-value for limit meta-analysis, t-value for all others; p-value: p-value of the effect of education; tau<sup>2</sup> / I<sup>2</sup>: measures of heterogeneity for main analysis, measures of residual heterogeneity for overall subgroup analyses, measures of heterogeneity within the subgroup for individual subgroups; Q/Q'-Q/df/p-value of Q = results of test of heterogeneity for main analysis and for individual subgroups, results of test for subgroup differences for overall subgroup analyses, results of test of small-study effects for limit meta-analysis df = degrees of freedom; HIC = high-income country, L/MIC = low- or middle-income country

**Supplementary Table 6.** Results of meta-analysis excluding outliers reporting other effect measures than odds ratios.

| Results of meta-analyses excluding studies reporting other effect measures than odds ratios |  |  |  |  |  |  |  |  |  |  |
| --- | --- | --- | --- | --- | --- | --- | --- | --- | --- | --- |
| Analysis | n studies | OR (95% CI) | t-/z-value | p-value | prediction interval | tau<sup>2</sup> | I<sup>2</sup> (%) | Q / Q'-Q | df | p-value of Q |
| main | 18 | 1.1 (0.87 - 1.38) | 0.866 | 0.398 | 0.46 - 2.61 | 0.158 | 76.5 | 72.332 | 17 | 0.000 |
| subgroup smoking | 18 | 1.1 (0.87 - 1.38) | 0.866 | 0.398 | 0.46 - 2.61 | 0.152 | 75.0 | 1.586 | 1 | 0.208 |
| smoking not controlled | 12 | 1 (0.74 - 1.36) | -0.007 | 0.995 | 0.4 - 2.47 | 0.152 | 72.0 | 39.321 | 16 | 0.000 |
| smoking controlled | 6 | 1.3 (0.87 - 1.94) | 1.680 | 0.154 | 0.43 - 3.91 | 0.152 | 79.8 | 24.739 |  | 0.000 |
| subgroup representative | 18 | 1.1 (0.87 - 1.38) | 0.866 | 0.398 | 0.46 - 2.61 | 0.131 | 76.2 | 3.141 | 1 | 0.076 |
| not representative | 9 | 0.9 (0.6 - 1.36) | -0.579 | 0.578 | 0.37 - 2.22 | 0.131 | 77.1 | 34.872 | 16 | 0.000 |
| representative | 9 | 1.3 (1.03 - 1.63) | 2.618 | 0.031 | 0.53 - 3.16 | 0.131 | 75.2 | 32.276 |  | 0.000 |
| subgroup PD assessment | 18 | 1.1 (0.87 - 1.38) | 0.866 | 0.398 | 0.46 - 2.61 | 0.167 | 77.9 | 0.406 | 1 | 0.524 |
| not all assessed | 11 | 1.05 (0.73 - 1.5) | 0.280 | 0.785 | 0.4 - 2.74 | 0.167 | 83.5 | 60.590 | 16 | 0.000 |
| all assessed | 7 | 1.19 (0.9 - 1.56) | 1.537 | 0.175 | 0.4 - 3.54 | 0.167 | 48.9 | 11.734 |  | 0.068 |
| subgroup income | 18 | 1.1 (0.87 - 1.38) | 0.866 | 0.398 | 0.46 - 2.61 | 0.167 | 77.5 | 0.367 | 1 | 0.545 |
| HIC | 7 | 1.19 (0.82 - 1.73) | 1.148 | 0.295 | 0.4 - 3.57 | 0.167 | 49.2 | 11.805 | 16 | 0.066 |
| L/MIC | 11 | 1.05 (0.75 - 1.46) | 0.308 | 0.764 | 0.4 - 2.74 | 0.167 | 83.1 | 59.196 |  | 0.000 |
| limit meta-analysis | 18 | 1.13 (0.8 - 0.8) | 0.697 | 0.486 |  | 0.158 | 76.5 | 0.213 | 1 | 0.645 |

OR (95% CI): Odds ratio with 95% confidence interval (adjusted for limit meta-analysis row); t/z-value: z-value for limit meta-analysis, t-value for all others; p-value: p-value of the effect of education; tau<sup>2</sup> / I<sup>2</sup>: measures of heterogeneity for main analysis, measures of residual heterogeneity for overall subgroup analyses, measures of heterogeneity within the subgroup for individual subgroups; Q/Q'-Q/df/p-value of Q = results of test of heterogeneity for main analysis and for individual subgroups, results of test for subgroup differences for overall subgroup analyses, results of test of small-study effects for limit meta-analysis df = degrees of freedom; HIC = high-income country, L/MIC = low- or middle-income country

**Supplementary Table 7.** Results of meta-analysis excluding mendelian randomization stuties.

| Results of meta-analyses excluding mendelian randomization studies |  |  |  |  |  |  |  |  |  |  |
| --- | --- | --- | --- | --- | --- | --- | --- | --- | --- | --- |
| Analysis | n studies | OR (95% CI) | t-/z-value | p-value | prediction interval | tau<sup>2</sup> | I<sup>2</sup> (%) | Q / Q'-Q | df | p-value of Q |
| main | 22 | 1.04 (0.88 - 1.22) | 0.497 | 0.624 | 0.54 - 2 | 0.094 | 74.0 | 80.870 | 21 | 0.000 |
| subgroup smoking | 22 | 1.04 (0.88 - 1.22) | 0.497 | 0.624 | 0.54 - 2 | 0.102 | 75.2 | 0.005 | 1 | 0.943 |
| smoking not controlled | 17 | 1.04 (0.85 - 1.27) | 0.440 | 0.666 | 0.52 - 2.1 | 0.102 | 76.6 | 68.442 | 20 | 0.000 |
| smoking controlled | 5 | 1.03 (0.68 - 1.56) | 0.191 | 0.858 | 0.38 - 2.8 | 0.102 | 67.6 | 12.363 |  | 0.015 |
| subgroup representative | 22 | 1.04 (0.88 - 1.22) | 0.497 | 0.624 | 0.54 - 2 | 0.087 | 67.6 | 1.656 | 1 | 0.198 |
| not representative | 10 | 0.91 (0.65 - 1.29) | -0.584 | 0.574 | 0.45 - 1.88 | 0.087 | 74.2 | 34.872 | 20 | 0.000 |
| representative | 12 | 1.13 (0.98 - 1.31) | 1.886 | 0.086 | 0.57 - 2.24 | 0.087 | 59.0 | 26.832 |  | 0.005 |
| subgroup PD assessment | 22 | 1.04 (0.88 - 1.22) | 0.497 | 0.624 | 0.54 - 2 | 0.087 | 73.3 | 2.501 | 1 | 0.114 |
| not all assessed | 14 | 0.97 (0.77 - 1.21) | -0.340 | 0.739 | 0.5 - 1.88 | 0.087 | 78.9 | 61.596 | 20 | 0.000 |
| all assessed | 8 | 1.21 (0.96 - 1.52) | 1.942 | 0.093 | 0.56 - 2.59 | 0.087 | 47.0 | 13.205 |  | 0.067 |
| subgroup income | 22 | 1.04 (0.88 - 1.22) | 0.497 | 0.624 | 0.54 - 2 | 0.085 | 71.5 | 2.397 | 1 | 0.122 |
| HIC | 8 | 1.23 (0.91 - 1.65) | 1.633 | 0.146 | 0.58 - 2.61 | 0.085 | 43.6 | 12.420 | 20 | 0.088 |
| L/MIC | 14 | 0.96 (0.79 - 1.18) | -0.400 | 0.696 | 0.5 - 1.86 | 0.085 | 77.5 | 57.732 |  | 0.000 |
| limit meta-analysis | 22 | 1.05 (0.84 - 0.84) | 0.439 | 0.661 |  | 0.094 | 74.0 | 4.059 | 1 | 0.044 |

OR (95% CI): Odds ratio with 95% confidence interval (adjusted for limit meta-analysis row); t/z-value: z-value for limit meta-analysis, t-value for all others; p-value: p-value of the effect of education; tau<sup>2</sup> / I<sup>2</sup>: measures of heterogeneity for main analysis, measures of residual heterogeneity for overall subgroup analyses, measures of heterogeneity within the subgroup for individual subgroups; Q/Q'-Q/df/p-value of Q = results of test of heterogeneity for main analysis and for individual subgroups, results of test for subgroup differences for overall subgroup analyses, results of test of small-study effects for limit meta-analysis df = degrees of freedom; HIC = high-income country, L/MIC = low- or middle-income country

**Supplementary Table 8.** Results of meta-analysis excluding studies with a high risk of bias.

| Results of meta-analyses excluding studies with a high risk of bias |  |  |  |  |  |  |  |  |  |  |
| --- | --- | --- | --- | --- | --- | --- | --- | --- | --- | --- |
| Analysis | n studies | OR (95% CI) | t-/z-value | p-value | prediction interval | tau<sup>2</sup> | I<sup>2</sup> (%) | Q / Q'-Q | df | p-value of Q |
| main | 13 | 1.05 (0.77 - 1.42) | 0.315 | 0.758 | 0.38 - 2.9 | 0.199 | 78.3 | 55.422 | 12 | 0.000 |
| subgroup smoking | 13 | 1.05 (0.77 - 1.42) | 0.315 | 0.758 | 0.38 - 2.9 | 0.224 | 80.2 | 0.009 | 1 | 0.924 |
| smoking not controlled | 8 | 1.03 (0.62 - 1.73) | 0.154 | 0.882 | 0.31 - 3.47 | 0.224 | 80.6 | 36.004 | 11 | 0.000 |
| smoking controlled | 5 | 1.06 (0.68 - 1.66) | 0.369 | 0.731 | 0.25 - 4.58 | 0.224 | 79.4 | 19.416 |  | 0.001 |
| subgroup representative | 13 | 1.05 (0.77 - 1.42) | 0.315 | 0.758 | 0.38 - 2.9 | 0.179 | 75.5 | 1.710 | 1 | 0.191 |
| not representative | 6 | 0.84 (0.42 - 1.68) | -0.651 | 0.544 | 0.25 - 2.81 | 0.179 | 80.9 | 26.216 | 11 | 0.000 |
| representative | 7 | 1.23 (0.93 - 1.62) | 1.835 | 0.116 | 0.4 - 3.78 | 0.179 | 67.9 | 18.686 |  | 0.005 |
| subgroup PD assessment | 13 | 1.05 (0.77 - 1.42) | 0.315 | 0.758 | 0.38 - 2.9 | 0.192 | 80.1 | 1.464 | 1 | 0.226 |
| not all assessed | 7 | 0.9 (0.51 - 1.6) | -0.448 | 0.670 | 0.28 - 2.9 | 0.192 | 86.4 | 44.034 | 11 | 0.000 |
| all assessed | 6 | 1.24 (0.9 - 1.7) | 1.748 | 0.141 | 0.36 - 4.29 | 0.192 | 55.7 | 11.293 |  | 0.046 |
| subgroup income | 13 | 1.05 (0.77 - 1.42) | 0.315 | 0.758 | 0.38 - 2.9 | 0.192 | 78.5 | 1.406 | 1 | 0.236 |
| HIC | 5 | 1.3 (0.71 - 2.36) | 1.198 | 0.297 | 0.33 - 5.14 | 0.192 | 56.5 | 9.191 | 11 | 0.057 |
| L/MIC | 8 | 0.93 (0.61 - 1.42) | -0.413 | 0.692 | 0.3 - 2.83 | 0.192 | 83.3 | 41.989 |  | 0.000 |
| limit meta-analysis | 13 | 1.02 (0.62 - 0.62) | 0.059 | 0.953 |  | 0.199 | 78.3 | 3.150 | 1 | 0.076 |

OR (95% CI): Odds ratio with 95% confidence interval (adjusted for limit meta-analysis row); t/z-value: z-value for limit meta-analysis, t-value for all others; p-value: p-value of the effect of education; tau<sup>2</sup> / I<sup>2</sup>: measures of heterogeneity for main analysis, measures of residual heterogeneity for overall subgroup analyses, measures of heterogeneity within the subgroup for individual subgroups; Q/Q'-Q/df/p-value of Q = results of test of heterogeneity for main analysis and for individual subgroups, results of test for subgroup differences for overall subgroup analyses, results of test of small-study effects for limit meta-analysis df = degrees of freedom; HIC = high-income country, L/MIC = low- or middle-income country

**Supplementary Table 9.** Results of meta-analysis including only one Swedish register based study.

| Results of meta-analyses including only one Swedish register-based study |  |  |  |  |  |  |  |  |  |  |
| --- | --- | --- | --- | --- | --- | --- | --- | --- | --- | --- |
| Analysis | n studies | OR (95% CI) | t-/z-value | p-value | prediction interval | tau<sup>2</sup> | I<sup>2</sup> (%) | Q / Q'-Q | df | p-value of Q |
| main | 22 | 1.06 (0.89 - 1.27) | 0.724 | 0.477 | 0.5 - 2.25 | 0.121 | 76.8 | 90.487 | 21 | 0.000 |
| subgroup smoking | 22 | 1.06 (0.89 - 1.27) | 0.724 | 0.477 | 0.5 - 2.25 | 0.126 | 75.4 | 0.504 | 1 | 0.478 |
| smoking not controlled | 16 | 1.02 (0.82 - 1.27) | 0.222 | 0.827 | 0.47 - 2.25 | 0.126 | 71.3 | 52.351 | 20 | 0.000 |
| smoking controlled | 6 | 1.18 (0.75 - 1.85) | 0.949 | 0.386 | 0.43 - 3.22 | 0.126 | 82.8 | 29.090 |  | 0.000 |
| subgroup representative | 22 | 1.06 (0.89 - 1.27) | 0.724 | 0.477 | 0.5 - 2.25 | 0.109 | 72.7 | 2.216 | 1 | 0.137 |
| not representative | 10 | 0.91 (0.64 - 1.3) | -0.594 | 0.567 | 0.41 - 2.03 | 0.109 | 74.2 | 34.872 | 20 | 0.000 |
| representative | 12 | 1.19 (0.98 - 1.43) | 2.007 | 0.070 | 0.55 - 2.55 | 0.109 | 71.3 | 38.363 |  | 0.000 |
| subgroup PD assessment | 22 | 1.06 (0.89 - 1.27) | 0.724 | 0.477 | 0.5 - 2.25 | 0.120 | 76.3 | 1.597 | 1 | 0.206 |
| not all assessed | 14 | 1 (0.77 - 1.29) | -0.037 | 0.971 | 0.46 - 2.18 | 0.120 | 81.7 | 71.033 | 20 | 0.000 |
| all assessed | 8 | 1.21 (0.96 - 1.52) | 1.963 | 0.090 | 0.5 - 2.95 | 0.120 | 47.0 | 13.205 |  | 0.067 |
| subgroup income | 22 | 1.06 (0.89 - 1.27) | 0.724 | 0.477 | 0.5 - 2.25 | 0.119 | 74.9 | 1.471 | 1 | 0.225 |
| HIC | 8 | 1.22 (0.9 - 1.66) | 1.561 | 0.163 | 0.5 - 2.97 | 0.119 | 43.6 | 12.420 | 20 | 0.088 |
| L/MIC | 14 | 0.99 (0.78 - 1.26) | -0.048 | 0.962 | 0.46 - 2.17 | 0.119 | 80.6 | 67.116 |  | 0.000 |
| limit meta-analysis | 22 | 1.07 (0.82 - 0.82) | 0.490 | 0.624 |  | 0.121 | 76.8 | 6.234 | 1 | 0.013 |

OR (95% CI): Odds ratio with 95% confidence interval (adjusted for limit meta-analysis row); t/z-value: z-value for limit meta-analysis, t-value for all others; p-value: p-value of the effect of education; tau<sup>2</sup> / I<sup>2</sup>: measures of heterogeneity for main analysis, measures of residual heterogeneity for overall subgroup analyses, measures of heterogeneity within the subgroup for individual subgroups; Q/Q'-Q/df/p-value of Q = results of test of heterogeneity for main analysis and for individual subgroups, results of test for subgroup differences for overall subgroup analyses, results of test of small-study effects for limit meta-analysis df = degrees of freedom; HIC = high-income country, L/MIC = low- or middle-income country

**Supplementary Table 10.** Results of the p-curve analysis of the meta-analysis excluding outliers.

|  |  |  |  |  |  |  |  |  |  |  |  |
| --- | --- | --- | --- | --- | --- | --- | --- | --- | --- | --- | --- |
| n of studies | n p < 0.05 | n p < 0.025 | Test | p binomial | z full | p full | z half | p half | Power estimate | Evidential value | Effect estimate |
| 22 | 6 | 4 | right-skewness | 0.344 | -2.711 | 0.003 | -2.861 | 0.002 | 0.59 (0.167 - 0.888) | yes | 0.008 |
| 22 | 6 | 4 | flatness | 0.549 | 0.952 | 0.829 | 3.055 | 0.999 | 0.59 (0.167 - 0.888) | no | 0.008 |

Power estimate: The power estimate and 95% confidence interval; Effect estimate: estimated true effect d; p binomial: p-value of the binomial test comparing significant p-values above and below 0.025; z/p full/half: effect measures and measures of significance of the pp-value based tests of right-skewness/flatness for the full sample (all significant p-values) and half of the sample (only p-values > 0.025)
The test of right-skewness tests for evidential for an actual effect. The test of flatness tests for evidential value of an absence of a real effect.

**Supplementary Table 11.**
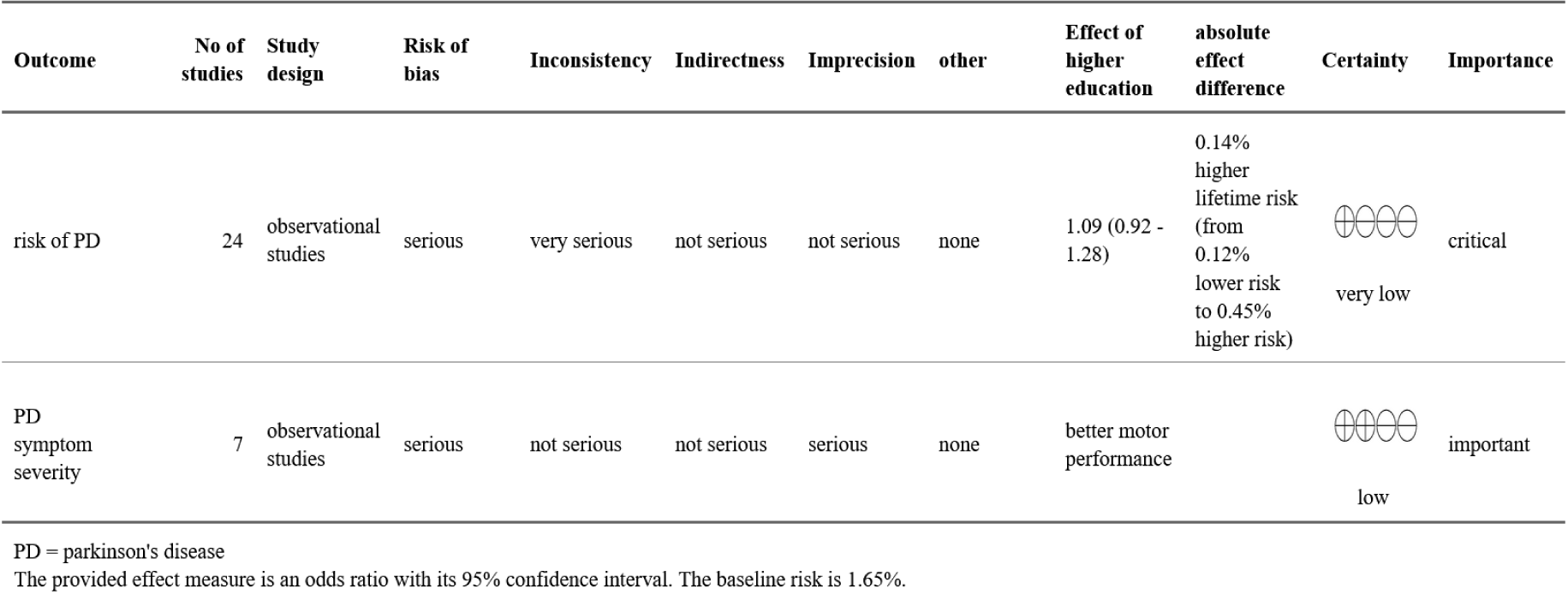
GRADE evidence profile.

| GRADE evidence profile |  |  |  |  |  |  |  |  |  |  |  |
| --- | --- | --- | --- | --- | --- | --- | --- | --- | --- | --- | --- |
| Outcome | No of studies | Study design | Risk of bias | Inconsistency | Indirectness | Imprecision | other | Effect of higher education | absolute effect difference | Certainty | Importance |
| risk of PD | 24 | observational studies | serious | very serious | not serious | not serious | none | 1.09 (0.92 - 1.28) | 0.14% higher lifetime risk (from 0.12% lower risk to 0.45% higher risk) | very low | critical |
| PD symptom severity | 7 | observational studies | serious | not serious | not serious | serious | none | better motor performance |  | low | important |

PD = parkinson's disease
The provided effect measure is an odds ratio with its 95% confidence interval. The baseline risk is 1.65%.

